# “Like taking part in Star Wars”: A thematic analysis of acceptability and experiences of older adults participating in remote longitudinal sleep and dementia research

**DOI:** 10.1101/2025.11.19.25340565

**Authors:** Bijetri Biswas, Victoria Grace Gabb, Jonathan Blackman, Hamish Morrison, Elizabeth Coulthard, Anne Roudaut

**Affiliations:** ReMemBr Group, North Bristol NHS Trust, Southmead Hospital, Bristol, United Kingdom, BS10 5NB; Faculty of Science and Engineering, University of Bristol, Bristol, United Kingdom, BS8 1TR; Bristol Medical School, University of Bristol, Bristol, United Kingdom, BS8 1UD; NIHR Bristol Biomedical Research Centre, Bristol, United Kingdom, BS2 8DX

**Keywords:** dementia, mild cognitive impairment, sleep, technology, digital health, wearables, acceptability, engagement, usability, patient-centred research

## Abstract

**Background:** Sleep disturbance is a common symptom of and potential risk factor for neurodegeneration. Remote sleep and cognitive assessments offer promise for monitoring symptoms and treatment response from patients’ homes, but the acceptability of remote sleep and circadian technology in older adults with and without cognitive impairment is not known.

**Objective:** This qualitative study was designed to explore and describe the barriers, facilitators, and user experience of older adults with mild cognitive impairment and dementia and cognitively unimpaired older adults who participated in a longitudinal sleep and memory study designed around remote monitoring technologies.

**Methods:** Patients with mild cognitive impairment or dementia due to probable Alzheimer’s disease or Lewy body disease and age-matched controls participated in a longitudinal remote study involving multimodal assessments of sleep and cognition including actigraphy, wireless electroencephalography, a smartphone app, web-based cognitive tasks, and serial saliva samples. Participants were asked for feedback via questionnaires during the study and invited to complete end-of-study interviews about their experiences. Questions were informed and thematic analysis was guided by the Capability, Opportunity, Motivation – Behaviour model of behaviour change and the extended Unified Theory of Acceptance and Use of Technology and focused on perceived barriers and facilitators.

**Results:** The study identified six key themes. The first theme, ‘motivations to participate’, highlighted how participants felt the research could be helpful to themselves and others. The second theme, ‘navigating the user experience of devices’, identified comfort, security, privacy, ease of use, and reliability as fundamental in determining acceptability. ‘Adjusting over time to study participation’, the third theme, covered changing perceptions with increased exposure and familiarity, and the importance of convenience, flexibility, and developing a routine. The fourth theme explored ‘social support as a facilitator and barrier to research participation’, looking at the influence of both the research team and relatives supporting at home. A fifth theme of ‘adherence, accuracy, and getting it right’ was also identified, as participants were motivated to provide good quality data for the study. Finally, we identified a sixth theme surrounding participants’ ‘reflections, realities, and uncertainties around sleep’, which focused on sleep hygiene and common sleeping problems in older adults, such as snoring and nocturnal awakenings.

**Conclusions:** Older adults with and without cognitive impairment were motivated to engage in longitudinal remote sleep research, follow remote research protocols, and produce good quality data. Acceptability was related to burden and convenience, usability, and emotional responses to study tasks. When study tasks are repeated over time, care should be taken to introduce variety where possible to avoid fatigue and frustration. Study partners offer essential support for some participants, but requiring a study partner may also be an unnecessary barrier to research participation for others. Future studies should aim to identify effective strategies for recruiting diverse populations, particularly those with limited technology experience or from underserved communities, to ensure equitable participation and representation in research. Providing education on the importance of sleep for brain health and technology use may be beneficial.

## Introduction

More than 55 million people live with dementia worldwide and many more have neurodegenerative diseases such as Alzheimer’s disease (AD) and Parkinson’s disease at earlier asymptomatic or mild cognitive impairment (MCI) stages ^1,2^. Recently, amyloid-targeting disease-modifying treatments have been shown to produce small clinical benefit in people with MCI or early dementia due to AD ^3^. However, most patients in memory clinics are not suitable for these new therapies ^4^ and novel therapeutic approaches are desperately needed ^5^.

### Sleep, ageing, and dementia

Nearly half of older adults report sleep complaints such as poor sleep quality, insomnia, and daytime fatigue ^6^. Sleep patterns undergo significant changes in older adulthood, including reduced sleep duration, increased sleep fragmentation, and dampened circadian rhythms ^7^. Common comorbidities associated with ageing including depression, nocturia, cardiovascular problems, and polypharmacy are also associated with sleep disturbance ^8^. However, sleep disturbance is also a common symptom of dementia and can occur early in the disease course. In AD, sleep disturbances often reflect an exaggeration of the changes observed in healthy ageing ^9^. People with Lewy body disease (LBD), which includes dementia with Lewy bodies and Parkinson’s disease dementia, often first present with rapid eye movement (REM) sleep behaviour disorder and tend to report more subjective sleep disturbances than people with AD ^10,11^.

In addition to being symptomatic of neurodegeneration, sleep is also a potentially modifiable risk factor for dementia. Persistent short sleep duration, less than six hours per night, is associated with increased dementia risk ^12^ and sleep disorders such as insomnia, idiopathic REM sleep behaviour disorder, and sleep apnoea have all been linked to increased risk of dementia ^13^. Proposed mechanisms through which sleep disturbances may contribute towards the risk of dementia include impairing cerebral clearance of metabolic waste products including amyloid-beta ^14^, increasing inflammation ^15^, and adversely impacting cardiovascular health ^16^. Given that sleep plays a crucial role in cognitive and brain health, therapeutic interventions to improve sleep could contribute to both prevention and treatment of dementia ^17^. However, we first need to better understand the mechanisms linking sleep and dementia and identify therapeutic targets relevant to different underlying aetiologies.

### Measuring sleep in individuals with and at risk of dementia

To date, sleep research in dementia or MCI has largely utilised self-report or polysomnography within a clinical or laboratory environment ^18^. Self-report measures can capture important data on subjective sleep quality, but have issues with recall bias, poorly align with objective measures of sleep, and cannot capture sleep microarchitecture or sleep staging ^19^. Some of the most commonly utilised questionnaires also ask participants to describe their typical recent sleep (e.g., ‘during the past month’ ^20^ or ‘in recent times’ ^21^), which can be difficult for people with cognitive impairment and may minimise important variation across nights. Whilst polysomnography gives much richer objective data, it is impractical in longitudinal or large-scale studies due to the resource needed, and the artificial environment of the clinic can be distressing and disrupts normal sleep-wake routines ^22^. Before we can invest in large-scale clinical trials of sleep, it is important to identify methods to measure sleep longitudinally in older adults in a scalable and acceptable way.

Remote monitoring technologies (RMTs), including wearables such as wireless electroencephalography (EEG) sleep headbands and actigraphy devices, offer alternatives to polysomnography and questionnaires which could be scalable, integrate into normal routines, and provide rich multimodal data ^23^. To complement RMTs, sleep researchers can utilise other remote study tasks, which can be supervised or unsupervised, to allow most if not all study data to be collected in the home environment, reducing the burden of lengthy clinic visits and capturing more naturalistic data. For example, sleep researchers can use salivary assays instead of serial blood sampling for cortisol and melatonin ^24^ and utilise frequent remote cognitive testing instead of traditional clinic-based cognitive assessments ^25,26^. In combination, digital health technologies could offer in-depth longitudinal sleep and cognitive profiling from home which might be more sensitive to changes over time, better capture intra-individual variation, and offer a pragmatic way to monitor the safety and efficacy of new treatments targeting sleep ^27^.

However, the acceptability of remote sleep and dementia research designs is not yet understood. Whilst many older adults and people living with MCI or dementia are now digitally proficient and using the internet and digital health technologies to support their daily lives ^28,29^, large numbers of older adults still have limited or no access to technology^30^. Technology can offer opportunities to engage in activities and support daily living, but is not always designed with inclusivity or accessibility in mind and can be stressful for older adults and those with cognitive difficulties ^31^.

Cognitive impairment may directly affect how someone interacts with technology, their confidence or ability to learn new processes such as how to use apps, or their overall user experience ^32^. Older adults, particularly those with motor symptoms such as in LBD or comorbid health conditions such as arthritis, may also have difficulty using technology as it often requires fine motor skills ^33^. Concerns about data privacy are also often raised among older adults when considering the risks and benefits of technology ^34^. Tailoring technology to the unique needs and preferences of older adults can significantly enhance engagement and compliance ^30^. Therefore, studies exploring the user experience of remote sleep and memory assessments in older adults with and at-risk of dementia are crucial for optimising these technologies before use in clinical trials, to ensure digital health technologies are inclusive and acceptable.

### Aims of the present study

We aimed to evaluate how older adults with and without MCI or dementia perceived and experienced remote research design using multimodal remote sleep and cognitive digital health technologies. Specifically, we aimed to explore acceptability, engagement, barriers, and facilitators among older adults who participated in a remote sleep and cognitive research study conducted over eight weeks. We also wanted to find out how much and what kind of study support might be needed to enable remote sleep and dementia research and how we could improve and inform future research design.

## Methods

### Overview

This analysis is part of a larger observational prospective cohort study called the Remote Evaluation of Sleep to Enhance Understanding of Early Dementia (RESTED) study, which was designed to examine the acceptability and feasibility of remote sleep monitoring in older adults with and without MCI and dementia ^35^ and compare sleep differences between the cohorts ^36^. Quantitative feasibility results (e.g., on adherence and data quality) have been published elsewhere^37^. Here, we focus on the acceptability of the remote study design for sleep and dementia research and draw on online interviews and questionnaires administered as part of the RESTED study in a thematic analysis. This study is reported in accordance with the Standards for Reporting Qualitative Research ^38^ (**Supplementary Materials**).

### Ethical considerations

This study was approved by the Health Research Authority (Yorkshire and the Humber—Bradford Leeds Research Ethics Committee, reference 21/YH/0177) and conducted in accordance with Good Clinical Practice and the Helsinki Declaration to protect the rights and welfare of all participants. All participants had capacity to consent and provided written informed consent before participation. Participants were reminded of their right to withdraw without it affecting their healthcare or opportunity to participate in other research studies. To protect participant anonymity, particularly given the sample size and the sensitive nature of the topic, identifiable data disclosed during interviews were removed and we chose not to associate quotes with individual identifiers. Instead, our analysis focused on shared thematic patterns, rather than individual-level variation. Quotes were selected to represent the breadth and diversity of perspectives expressed during the interviews.

### Reflexivity

Interviews were conducted by the joint first authors (BB and VG). BB has a background in digital health technology and VG predominantly works in sleep and dementia research, with a background in health research and neuroscience. As BB was primarily involved in the qualitative elements of the study, she was not known to participants prior to interviews, whereas as the lead research assistant and main point of contact for participants during the study, VG, was known to all research participants from consent onwards. This established familiarity was advantageous in fostering openness and trust during interviews but required ongoing reflexive awareness of how prior interactions might influence participants’ responses and the interpretation of their narratives. The authors made conscious efforts to discuss and challenge assumptions and perspectives that may have influenced their interpretation of the data. Reflexive memos were maintained throughout the analytic process to document interpretive decisions, challenge disciplinary biases, and support transparency during theme development.

Several steps were taken to diminish the distance or power imbalance in the ‘researcher-researched’ relationship ^39^. Interviews were conducted remotely with participants taking part from home at a time of their choosing, with partners present where participants felt more comfortable with this set-up ^40^. Conducting interviews remotely also allowed participants to engage from familiar surroundings, reducing potential anxiety associated with clinical environments and empowering them to control the timing and setting of participation. Within the interview, participants were actively encouraged to share both positive and negative experiences honestly to help the research team learn from them and share their advice and perspectives for future research. The semi-structured interview design and free-text responses in the questionnaire also allowed participants to guide conversations or comments towards what they perceived as important or relevant. These reflexive and participant-centred strategies aimed to strengthen the study’s credibility and trustworthiness by promoting openness, inclusion, and co-construction of meaning between researchers and participants.

### Participants

Of the 40 participants who completed the RESTED study, 32 (80%) participants provided data for the acceptability study through completing the end of study interview, questionnaire, or both. Characteristics of the participants included in this analysis are presented in Error! Reference source not found..

All participants met established clinical criteria for MCI or dementia due to probable AD or LBD or were age-matched cognitively healthy controls identified through cognitive and movement disorders clinics at North Bristol NHS Trust and research volunteer databases. Diagnoses of patients with MCI/dementia included in the present study included AD dementia (N=2), amnestic MCI or MCI due to AD (N=3), primary progressive aphasia due to probable AD (N=1), MCI due to LBD (N=4), and LBD dementia (N=3). Eligibility criteria required participants to be aged ≥ 50 years old, score ≥ 11 on the Montreal Cognitive Assessment (MoCA) during screening ^41^ and have no significant untreated comorbidities unrelated to an MCI/dementia diagnosis that could significantly interfere with sleep.

**Table 1.**
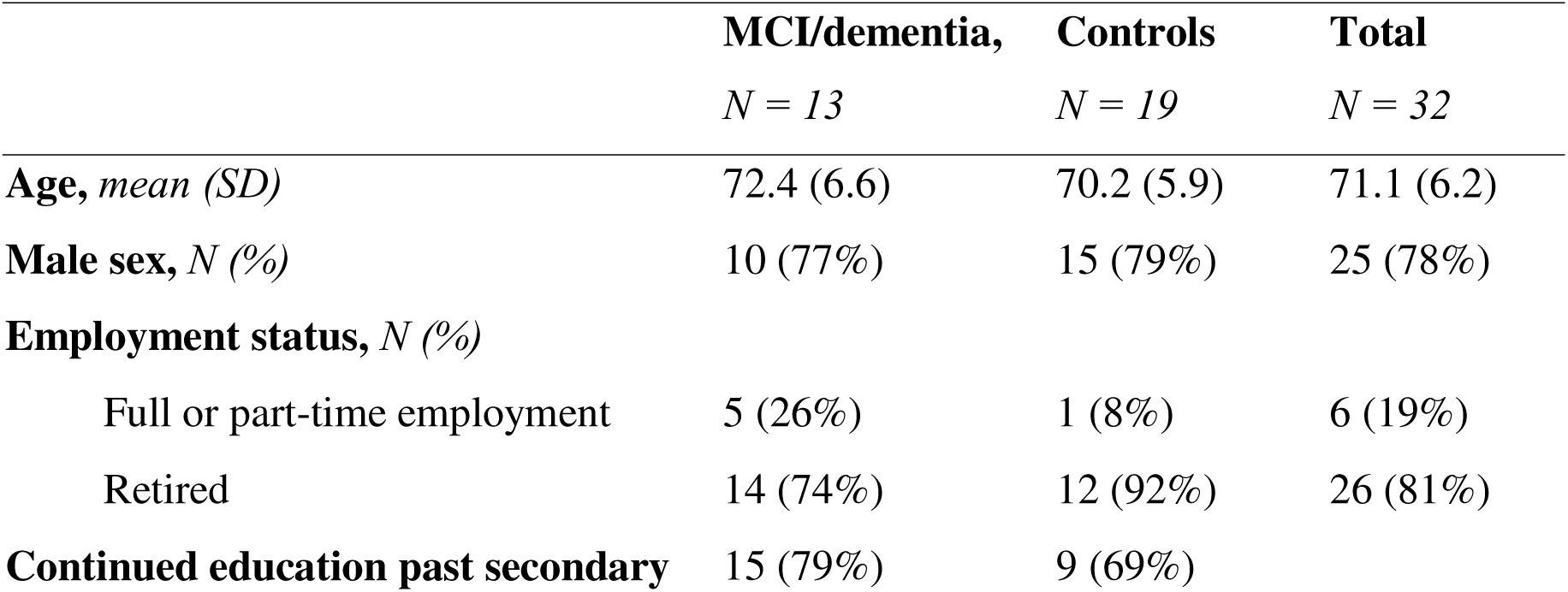
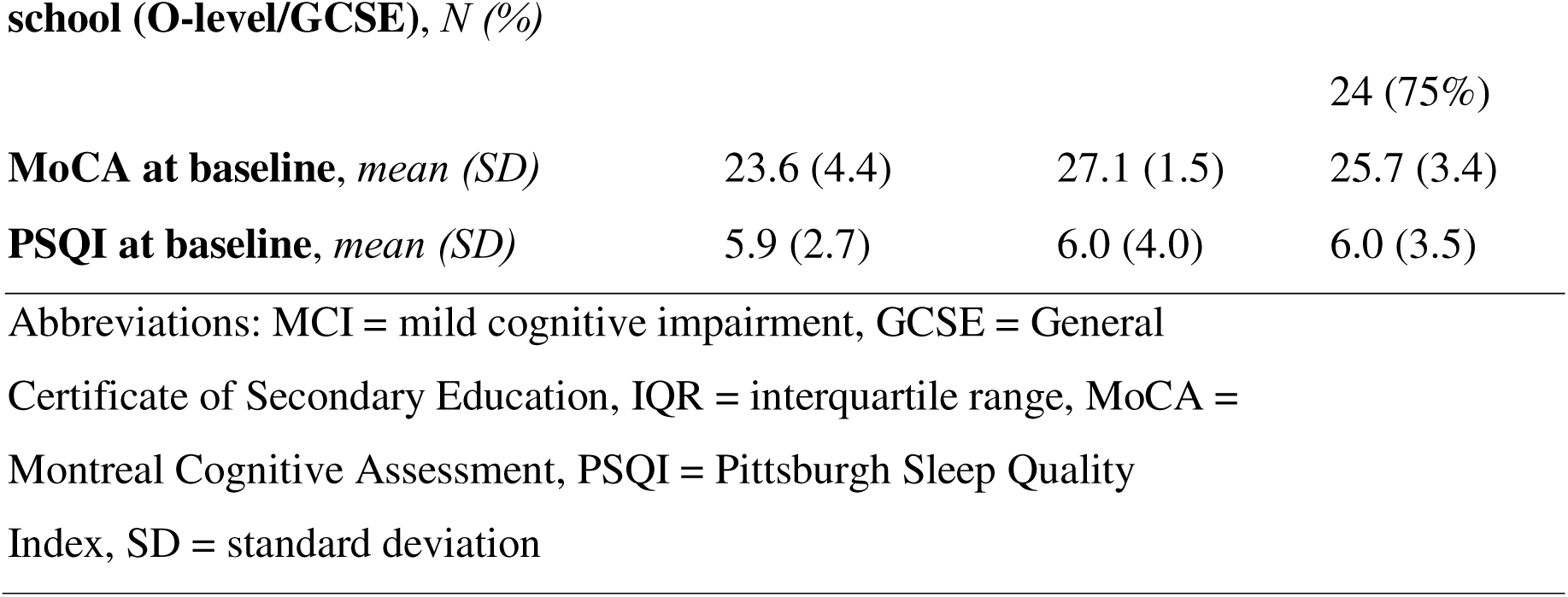
Baseline characteristics for participants included in the acceptability analysis.

### Study design

The full study protocol has been published online ^37^. Briefly, the RESTED study involved longitudinal remote monitoring and assessment of sleep and cognition at home whilst participants continued their usual activities. Following consent and screening, participants completed baseline assessments on sleep, cognition, and mood ^37^. For eight weeks during the main study period, objective sleep was assessed using a wrist-worn actigraphy device (*Axivity AX3*). Actigraphy data was complemented by daily digital sleep diaries collected via an app (*MyDignio*) installed on a mobile or tablet device. Twice weekly, participants completed remote unsupervised cognitive assessments via an online platform (*Cognitron*).

For 7 days during the main study period (the ‘intensive week’), additional data was collected. Objective sleep was measured using a wireless electroencephalography (EEG) headband (*Dreem 2*), which has been found to provide acceptable estimates of sleep parameters in this population ^42–44^. Cognition was assessed with daily unsupervised cognitive assessments (*Cognitron*) and two supervised verbal memory tests administered remotely via videoconferencing with the research team (*Microsoft Teams*) across two evenings to learn the word list and the following mornings to test recall and recognition. Circadian rhythms were measured using serial saliva samples collected at home to measure dim-light melatonin onset and cortisol awakening response across one evening and one morning respectively. Participants without a diagnosis of sleep apnoea or recent sleep apnoea assessment were also asked to complete two consecutive nights of overnight pulse oximetry (*Nonin 3150 WristOx2*). This combination of wearable devices, biological sampling, and digital tools aimed to offer a holistic and ecologically valid approach to studying sleep and cognition in individuals with MCI and dementia ^42^.

Figure 1 illustrates the study design and highlights when and how qualitative data was collected. Participants who were recruited to the RESTED study were invited to complete app-based questionnaires at the start of the study and following the intensive week and complete an end of study semi-structured interview. Interviews were conducted online and audio-recorded with verbal consent and typically lasted 30 to 60 minutes. The interview topic guide and questionnaires are provided in Error! Reference source not found..

**Figure 1.**
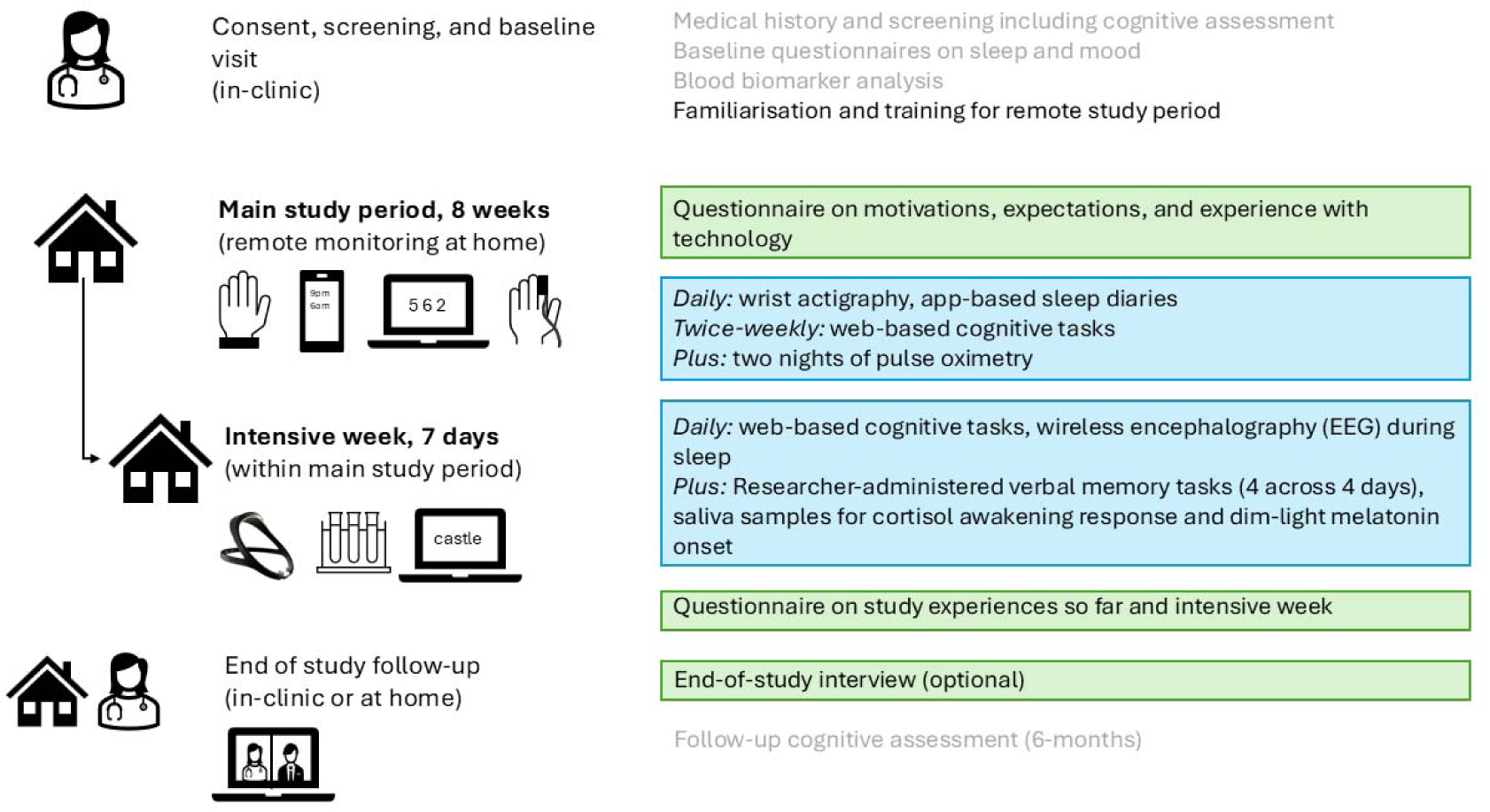
Overview of data collection in the Remote Evaluation of Sleep to Enhance Understanding of Early Dementia (RESTED) study. Remote study tasks were administered across eight weeks and are highlighted (blue). Qualitative data was collected during the main study period through questionnaires and an optional end of study interview (green). Study procedures in black text were the primary focus of this acceptability analysis.

### Data analysis

Audio recordings from online interviews were transcribed and pseudonymised using NVivo 12 software. Transcripts were coded by two authors (BB, VG) using Braun and Clarke’s phases of thematic analysis ^45^. The authors familiarised themselves with the data via independently reviewing each transcript and making initial notes. Preliminary codes were proposed, discussed, iteratively combined into broader initial themes before discussion and review with the wider study team. BB conducted a final check to ensure all relevant data were captured and themes were refined to reflect participants’ experiences accurately. Findings are presented below with detailed descriptions of the themes alongside illustrative quotes for each theme and sub-theme. Free-text questionnaire responses were also coded and integrated with the interview data to create a unified analysis.

A combination of deductive and inductive thematic analysis was conducted ^45^. Inductive coding allows themes to emerge directly from the data, whilst deductive coding complemented the research questions and allowed for integration of established theoretical approaches of understanding behaviour ^46^. The topic guide development, questionnaires, and later analysis was informed by the extended Unified Theory of Acceptance and Use of Technology (UTAUT-2)^47^ and the Capability, Opportunity, Motivation – Behaviour (COM-B)^48^ models. UTAUT-2 highlights how perceived benefits of technology (*performance expectancy*), ease (*effort expectancy*), *social influence* from important others, resources and support (*facilitating conditions*), fun (*hedonic motivation*), prior use (*habit)*, and *price value* influence technology acceptance ^47^ and how perceptions can be modified by age, gender, and experience. The COM-B model focuses on how knowledge, skills, and abilities (*capability*), external factors (*opportunity*), and internal processes (*motivation*) shape behaviour change, and can be applied to research engagement. Used together, these frameworks offered a complementary lens to explore both perceptions of technology and behavioural drivers of participation. The interconnection between these models and the themes is also discussed.

## Results

### Summary of themes

The thematic analysis resulted in five major themes around barriers and facilitators to engaging in remote longitudinal sleep and cognitive research: (1) motivations to participate; (2) navigating the user experience of devices; (3) adjusting to study participation over time; (4) social support as a facilitator and a barrier; (5) adherence, accuracy, and getting it right. The analysis also identified a sixth theme on reflections, realities, and uncertainties around sleep which captured older adults’ priorities and understanding of sleep. An overview of the themes is presented in Figure 2 with the content of themes presented in detail in the following sections.

**Figure 2.**
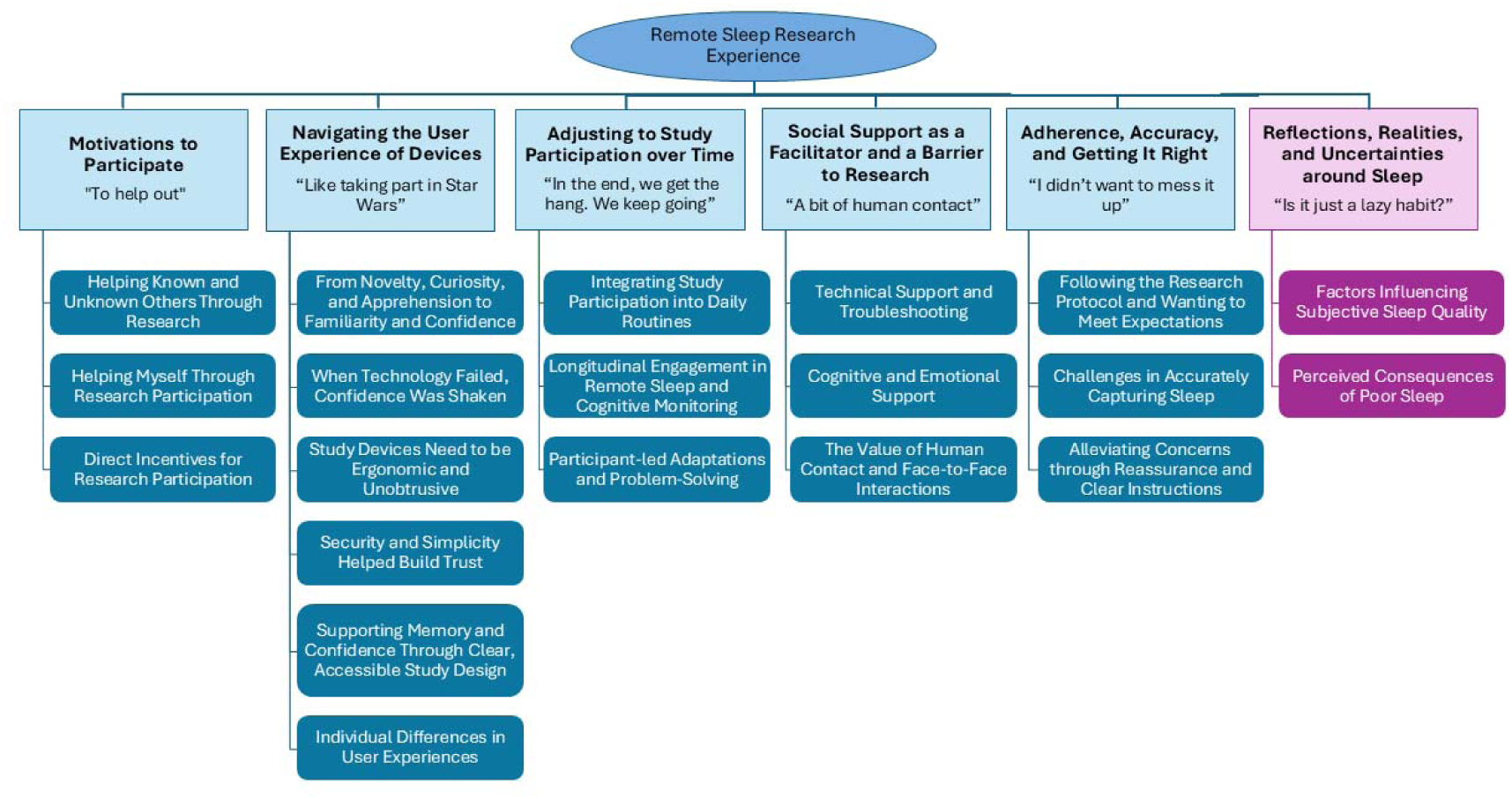
Overview of the results from the thematic analysis, capturing the key themes and their sub-themes. Five themes relating to study experiences and acceptability were identified (blue), with a sixth theme summarising participants’ understanding of sleep (purple).

#### Theme 1 – Motivations to Participate

“To help out”

##### Theme Overview

Participants predominantly reported taking part in the study because they perceived it as helpful either to themselves, family members who were at risk of dementia, or more widely to people with or at-risk of dementia. However, participants identified that additional incentives, such as providing feedback on sleep or financial reimbursement for time spent in the study, could also help to motivate more people to participate in future studies.

##### Helping Known and Unknown Others Through Research

A central motivation for participants was wanting to contribute towards research into dementia, as it could translate into better understanding of the condition and towards future treatments. Generally, participants wanted to:

> **“to support research into prevention, risk factors for, and management of, dementia”.**

Often, the underlying motivation was altruistic. Participants recognised that their contribution would likely not directly impact themselves, but might benefit others. Among older adults with a family history of dementia, participants hoped that participating might help family members who might develop dementia in the future, by helping to identify potentially modifiable risk factors and treatments:

> **“My two sisters and I are really quite concerned that we might get it […] it might not help me, but it might help my family further down the line.”**

Participants also shared sentiments of wanting to help the research team or clinic itself. One participant expressed how their appreciation for the service they receive from the healthcare and research team prompted research engagement:

> **“I get good treatment from [the clinic] and it is good to give something back.”**

##### Helping Myself Through Research Participation

However, other participants perceived direct personal benefits to participating in the research study. Study participation provided cognitive stimulation, which some participants felt might help delay cognitive decline or help to manage symptoms. As one participant explained:

> **“This sort of activity helps with my Alzheimer’s”.**

Participants also commented that participating might indicate if they needed further investigations for their cognition or sleep. One participant took part both for altruistic reasons, and to help them decide whether they needed additional support:

> **“In the hope it will help others in the future and hopefully give an indication whether I need to seek out help.”**

Being involved in research was also identified as a way to access potential treatments before they are approved:

> **“An idea that was suggested to me that that if anything is going on with Alzheimer’s and you’re in a research group, you might be the first to get something”.**

Positive affect was also a motivation for participating in the study. Several participants explained that they were took part in the study because it sounded **“interesting”** or that it would be something they might enjoy. Engaging in the study also helped participants to **“feel useful and part of something significant”.** Participants reported feeling **“good to contribute”**. Some participants also enjoyed having tasks to complete, as research gave them a sense of purpose or became a pastime:

> **“Having the structure and sort of something to be getting on with… I enjoyed it. Because once again, it gave me something to do.”**

Additionally, prior experience in research also appeared to facilitate participation, with several participants noting they took part because they had **“taken part in dementia studies before,”** suggesting research involvement can become habitual.

##### Direct Incentives for Research Participation

Although participants’ own motivations for participating centred around helping themselves or others, several suggested that adapting the study design or providing reimbursement to provide additional extrinsic motivation to participate could also be helpful. Offering direct incentives was considered as motivational for participating in research studies and a way to encourage more people to join research studies:

> **“It would encourage people, you know, to think that they were actually getting something out of it, rather than just entirely giving”.**

Several participants expressed an interest in receiving feedback on their cognitive performance and sleep data. Personalised feedback was seen as motivating, and could be incorporated into the design of RMTs, like screens on a smartwatch:

> **“If you could show me a data screen… I’d find that interesting.”**

Others requested more detailed or tangible outputs based on results, which could serve as a reminder or ‘thank you’ from the study:

> **· “I would love to have printouts of the graphs from the headset readings from all seven days to display on a wall”.**

Feedback on sleep was not provided to reduce the possibility of participants changing their behaviour in response to awareness of it. However, most participants who were interested in feedback did not intend to change their sleeping habits based on the feedback, unless the study identified a problem with their sleep such as an undiagnosed sleep disorder:

> **“If I was getting a lot of disturbed sleep and then yeah, it would be useful to know that. If you know, particularly if I was not picking up waking up in the night for example”.**

#### Theme 2 – Navigating the User Experience of Devices

“Like taking part in Star Wars”

##### Theme Overview

Participants’ first impressions of study devices were shaped by novelty and usability. User experience was influenced by factors such as ease of use, comfort, technical reliability, and privacy concerns, with digital literacy, age, cognitive impairment, and comorbidities likely affecting perceptions of usability.

##### From Novelty, Curiosity, and Apprehension to Familiarity and Confidence

Despite reporting frequent exposure to smart technology or familiarity with video calls at baseline, most study tasks were novel to participants. When asked about their first impressions, one participant remarked it was **“like taking part in Star Wars”** with another describing the technology as **“fascinating”,** suggesting that the study devices felt technologically advanced, exciting, or innovative.

Participants had mixed first impressions about the usability of devices. Participants often linked the unfamiliar study devices to familiar ones, creating mental anchors that helped them understand and contextualise their purpose. For example, the actigraphy devices were compared to smart fitness watches:

> **“I don’t even have, you know, a Fitbit or anything like that normally. So no, I hadn’t [come across the technology before]. Apps obviously are all a bit new.”**

Whilst some participants initially expressed feeling **“unsure”** or even **“daunted”** by the technology, others perceived devices as **“simple”** or **“straightforward”**. Effort expectancy was linked to digital literacy and confidence with technology, both of which improved as participants became more familiar with the devices.

For instance, one participant shared:

> **“At first, it didn’t appeal to me because I’m not very good with this sort of thing… [but] I haven’t had any problems. So, I must be better than I thought.”**

Changes in attitudes across the study is explored more in *Theme 3*.

##### When Technology Failed, Confidence Was Shaken

A key element in participants’ user experience of the study devices was *technical reliability,* or how consistently the technology performed according to participants’ expectations and aligned with the instructions provided:

> **“It was perfectly useful tech, you know, when it worked.”**

Several participants experienced technical glitches, predominantly triggered by software updates to the mobile application for the sleep diary and the website hosting the online cognitive tests. The software updates tended to affect whether a participant’s login details worked or stored a previous day’s data instead of allowing participants to enter new data. When expectations of the technology were not met, participants became frustrated:

> **“It didn’t signpost me to the Cognitron tests sometimes. It frequently told me […] I’d filled in the sleep diary and done the Cognitron tests when I hadn’t done either. That just seemed a bit irritating”.**

Participants were motivated to complete tasks, and sometimes blamed or questioned themselves when technology did not perform reliably:

> **“I thought it was me.”**

Often, participants were able to seek help from a partner or from the study team to find solutions or temporary workarounds, such as accessing the links on different web browsers whilst the research team contacted the developers to resolve the issues. However, in the meantime, technical glitches created additional participant burden and meant that participants were doubting themselves or becoming frustrated with the technology:

> **“I got my husband to look at the computer. I’m quite computer literate. And I got him to look at it and I said, ‘I don’t know what I’m doing wrong’ and he said, ‘I don’t think you’re doing anything wrong. It is a software problem’.”**

In addition to technical glitches, participants sometimes experienced confusion with interacting with different platforms and applications or websites for different tasks. Developing a single platform for all remote study tasks was suggested to make the study more straightforward for participants to keep track and minimise the need to familiarise themselves with different platforms:

> **“If there’s something that could link it all up in one app or one program”.**

##### Study Devices Need to be Ergonomic and Unobtrusive

Comfort and fit were often mentioned and could be considered a potential barrier to adoption of RMTs. For the most part, participants found the devices tolerable and that they did not interfere too much with their day-to-day life or their sleep:

> **“I found it very comfortable and not at all intrusive”.**

However, several participants suggested improvements to the actigraphy device and its strap. Continuous wear occasionally caused skin irritation, which was usually resolved by swapping wrists or cleaning the skin:

> **“To start with, I got a slight irritation… but actually it cleared up pretty quickly.”**

The strap material of the actigraph also tended to catch on clothing during daily tasks, which could interfere with activities of daily living like getting dressed:

> **“What the material is made of, it’s not slippery enough… you’d have to change the design a little bit.”**

Participants recommended making the device smaller, lighter, and more streamlined, similar to modern smartwatch design:

> **“It just seemed like… wearing a miniature brick.”**

Participants also often had concerns about the *Dreem 2* EEG headband, mainly that it might shift during sleep and interfere with both sleep quality and data accuracy. Some participants made efforts to adjust the fit to keep it in place and ensure reliable data collection, whilst others had fewer problems than anticipated:

> **“I was pleasantly surprised. Actually, I thought it was gonna keep falling off… I really didn’t imagine that was gonna stay on my head overnight. And it never came off. So I was absolutely astonished really.”**

The small red light on the *Dreem 2*, which indicated that the recording was in progress, was mentioned by several participants as disruptive to sleep, highlighting how small design features can substantially impact user experience and may contribute towards observer effects:

> **“If that red light could be put in a different place where you can’t see it, it would be much better because you know, I was meant to be trying to sleep better and it was making me keep waking up.”**

The relationship between study participation and continuing vs disrupting participants’ routines is explored further in *Theme 3*.

Finally, some patients felt that whilst individual devices were comfortable or tolerable, wearing too many study devices simultaneously became uncomfortable and potentially disruptive to sleep:

> **“Having the headband, wristband, and the pulse oximeter on at the same time, [I] was challenged by that.”**

##### Security and Simplicity Helped Build Trust

Privacy and data security were key to engagement, with many participants emphasising the importance of non-intrusive data collection, such as the absence of cameras or questions about sensitive topics like dream content:

> **“The idea of perhaps cameras on you, that would be horrible as well, wouldn’t it? It was that, if somebody was watching what you do when you’re asleep? Yeah, then you might do ridiculous things or not very dignified things. I’d feel that would be an invasion of privacy.”.**

Participants often described how study tasks were not perceived as invasive or intrusive and understood that their participation was voluntary, which fostered a sense of control:

> **“I think anything that I’ve been asked to do, you always get the option to refuse if you’re not comfortable with it. … So no, I didn’t find anything intrusive.”**

Protecting data security while keeping device and software interactions straightforward was also a priority, helping participants feel comfortable and empowered to participate:

> **“Getting that balance between keeping it simple, but also secure. And the participants who are of a certain age group, I guess, need that confidence and the simplicity”.**

##### Supporting Memory and Confidence Through Clear, Accessible Study Design

Participants with MCI or dementia were often concerned, particularly at the start of the study, whether their memory might impact participation. However, most participants remembered to complete tasks ^37^, either independently or following reminders sent by the research team:

> **“It was just, sort of, a slight kind of apprehension about being able to remember the actual tasks, but actually it didn’t, it wasn’t too bad”.**

Having clear printed instructions in study guides reassured participants and allowed them to problem solve whenever they needed it, rather than relying on memory recall for the study tasks:

> **“I was very happy to do it myself because the instructions were good and obviously the issue is me to being able to hold that information in my own brain between reading it and then actually having to do the test.”**

Instructions were also provided on a screen at the start of each set of the unsupervised cognitive tasks. This format was disliked by some participants as their difficulties with memory meant that they might not remember the instructions when engaging in a task:

> **“Well, you don’t then get access back to the instructions. So, you just get fed up.”**

There was also a reflection that instructions could be better complemented by more visual or practical demonstrations. Participants were talked through and shown how to use the study kit at the start and were offered a call with a researcher before the intensive week with additional study tasks. However, opportunities to physically practice tasks in the presence of the research team were also recommended to check participants’ understanding and improve learning on novel tasks such as the saliva samples:

> **“Despite having the instructions and everything, I would have found it helpful if…you showed me exactly how you did it, and I did it at the same time. Because honestly, I couldn’t really get how it was supposed to work”.**

##### Individual Differences in User Experiences

Participants often had different opinions on which study tasks were the easiest or the hardest to complete. For example, many participants found it particularly challenging to collect saliva samples at home, and reported both cognitive difficulties (understanding the instructions, accurately timing sample collection) and practical difficulties (fitting samples in around existing commitments and physically producing sufficient saliva):

> **“Those wretched saliva tests are an absolute nightmare. I have never in my life ever done a saliva test. Yeah, so parcelling out exactly what you’ve got to do, and then also realising how often you’ve got to do it, and then juggling the idea of what, how do you fit in your normal thing[s]?”**

However, other participants felt home saliva testing was straightforward, or even an enjoyable part of the study, highlighting how user experience can vary substantially across participants and be influenced by understanding and personal satisfaction:

> **“I enjoyed the physical thing, where physical things are involved as well as just the questions and answers. Like the saliva testing and things like that”.**

Similarly, participants expressed divergent preferences regarding whether it was preferable to have more tasks to do each day or have minimal requirements on most days alongside a few more intensive days. Some participants shared that they would rather have a single week of planned and more intensive data collection than additional tasks throughout the study duration, as this made planning easier:

> **“I think, actually, having that concentrated week is probably the best way to do it. That was the only thing that kind of slightly made it difficult – to identify a week where I could really keep it clear and prioritise. … I prefer that structure to, yeah, spreading it out”.**

However, other participants found this structure **“too intensive”** and **“very trying”** and would have preferred spreading out the additional tasks across the full study period to mean less time burden or disruption per day.

In terms of what impacted different user experiences, age was reflected upon. Participants suggested that as older adults may not be as familiar with technology and may struggle to learn it as they get older or experience cognitive impairment, advanced age may be a potential barrier to using RMTs:

> **“I’d imagine somebody a lot older would probably struggle with some of that, particularly the technology side of it”.**

Dexterity and other physical challenges, such as arthritis or parkinsonism, also complicated interactions with devices and study tasks. For example, responding quickly and accurately in the online cognitive tasks was complicated by both dementia-related conditions, such as tremor and dystonia, and unrelated comorbidities, such as arthritis:

> **“Partly because I’ve got arthritis in my fingers, partly because I’ve had […] an operation on this finger.”**

There were also physical challenges associated with wearing multiple devices at once. Participants with sleep apnoea who were using a continuous positive airway pressure (CPAP) mask had some difficulties with fitting the mask and the EEG headband comfortably:

> **“It was a little bit more awkward than it should have been, simply because I had the CPAP mask as well”.**

#### Theme 3 – Adjusting to Study Participation over Time

“In the end, we get the hang. We keep going.”

##### Theme Overview

Prolonged engagement in research encouraged participants to find ways of adapting to novel tasks and study requirements. Being able to integrate study tasks conveniently into existing routines or commitments facilitated engagement and reduced perceptions of burden, whilst time restrictions were a barrier, particularly for those with more demanding schedules. Perceptions of the study and its tasks were not static and tended to evolve over time.

##### Integrating Study Participation into Daily Routines

Being able to integrate study tasks within existing routines or commitments was a key facilitator to participant engagement and adherence. At the outset, some participants were concerned that study participation might be burdensome or disruptive to their daily lives. Concerns around time burden required to complete tasks daily and at specific times were prevalent:

> **“[What] I didn’t want it to do was to interfere with, you know, sort of my own life too much, and it didn’t… That was really my own concern, because we’re sort of busy people.”**

Being able to develop habits or routines to complete their study tasks around their usual activities helped to reduce participant burden:

> **“As soon as I woke up, get a cup of coffee, do the morning, some pieces, and then it was forgotten for the rest of the day”.**

The passive data collection enabled by the actigraphy watch also supported participants to continue with their usual activities as much as possible:

> **“I just put it on and left it on and it was fine. Didn’t really trouble me.”**

To minimise the burden associated with more intensive periods of data collection, some participants planned out the day and prepared ahead of time by placing equipment or devices required for study tasks in plain sight, highlighting the importance of executive functioning in managing study tasks:

> **“I chose to watch stuff I pre-recorded so I could pause it and do the saliva sample or whatever, then come back to it. So, I kind of managed it. Yeah. And the next morning just had the saliva kits near the bed and did the sleep diary. And then did those before breakfast and after breakfast. Whatever it was, just, just pre-planned it all and executed the plan”.**

To allow participants to integrate study tasks into their daily routines, convenience and flexibility was important. Using smartphones and digital tablets, compared to using pen and paper assessments or personal computers, enabled greater flexibility around when and where study tasks were completed, particularly for people who had busy schedules, such as those still working:

> **“The iPad makes doing the Cognitron test really accessible, doesn’t it? Because I can always do it on the move… Whereas with a laptop… I’d have to drag it down and get power supply plugged in and sit somewhere.”**

Tasks which took longer to do or were requested at specific times were more difficult to incorporate within established routines and created more burden for participants. For example, saliva sampling was often considered challenging because it took more time, and the short intervals repeatedly disrupted usual activities across the evening:

> **“The trickiest bit by far in terms of fitting things in was the saliva testing and fitting in food, especially because like lots of older people, I have, you know, other health conditions… an eating-related health condition I have is my IBS [irritable bowel syndrome] and you know, needing to eat little and often is part of it.”**

Participants generally accepted the inconvenience of time-limited tasks which occurred infrequently (e.g., once or twice during the study period). However, time-specific tasks which were repeated more frequently, such as completing web-based cognitive tests in the morning, were less acceptable. When unavailable during the designated time, some participants felt stressed or skipped the tasks altogether:

> **“It was demanding for me, because I have commitments on nearly every day of the week, which require me to get out of the house quite early.”**

In contrast, others adapted the tasks to better suit their routines. For instance, one participant spread the cognitive tests throughout the day instead of completing them in a single session:

> **“What I realised later on in that test is that I didn’t have to do them all together at the same time.”**

##### Longitudinal Engagement in Remote Sleep and Cognitive Monitoring

Participants’ attitudes towards the study tasks were often not static and changed over the course of the study.

Due to the number of study tasks and novel RMTs, there was often a learning curve involved in the study and understanding both how to complete tasks and how to fit them around existing commitments or routines. Despite early concerns, most participants adapted quickly:

> **“I very quickly got used to what was going on and how it was being done.”**

With time and experience, participants became more confident and were able to complete various tasks at differing time intervals without too much difficulty:

> **“I think once you started getting the hang of putting it all together, it was easier.”**

Participants’ perceptions of study tasks were dynamic and changed throughout the course of the study. However, familiarity and repetition could be both facilitators and barriers to engagement. For example, repeating cognitive tasks at regular intervals across the study period reduced the stress and allowed for familiarisation and learning:

> **“It took me a couple of goes before I was really confident that it was all OK and I could do it.”**

Gamification of the task helped participants to stay engage in repeated cognitive tasks, as some participants competed to improve own their previous score. When asked if they minded completing the same tasks or would have liked them to be less frequent, one participant shared:

> **“Never, didn’t mind doing them. No, I don’t think it made any difference. I got a bit competitive with it”.**

Some participants even self-reported benefits of regular cognitive testing beyond scope of the study, such as boosted self-esteem, improved subjective cognitive functioning, and enjoyment:

> **“Well, it made me feel very good when I found towards the end, I was getting better at it. I enjoyed it.”**

However, repeating the same study tasks over time could also lead to fatigue and a desire to discontinue with the study or certain tasks. Cognitive tasks could highlight participants’ cognitive concerns, meaning they often became more frustrated, anxious, or disheartened with greater exposure to the tasks:

> **“Towards the end of the week, I was getting stressed with it. I think because it starts to put in your mind that maybe you are going ‘doolally’. Maybe at the beginning you’re a lot more aware of the numbers and you’re concentrating harder.”**

Tasks which became progressively difficult appeared to prompt disengagement, even in participants who were initially enthusiastic about the tasks. Feeling demotivated may have meant participants started to put in less effort or performed worse over time:

> **“To start with, you know, I, sort of the competitive bit of me thought ‘right, you know – I’m going to find a way of, not defeating this thing exactly, but I am going to try really hard to improve my scores’. … But after a while, I kind of felt no matter how hard I tried, I just couldn’t. … I didn’t exactly give up, but I certainly didn’t try as hard.”**

Whilst participants understood the rationale behind repeating the same series of cognitive tasks was to enable comparison over time, the lack of variety meant that some participants transitioned from finding the study tasks engaging and enjoyable to tedious:

> **“I found it a chore doing them after the novelty had worn off, yeah. First, I thought, ‘Oh yes, I like puzzles.’ And then I thought, ‘no, not this puzzle’.”**

Despite challenges, participants’ sense of commitment and responsibility to the study encouraged them to keep going, despite experiencing study fatigue:

> **“I’m doing this for research purposes. There’s a reason why I’m doing it and it has to be done. I mean, you know, from the outset, you need to do it for that amount of time for it to be worthwhile.”**

##### Participant-led Adaptations and Problem-Solving

Throughout the study, participants often took initiative to make it easier to complete study tasks or improve their performance. Several participants developed strategies to help improve their cognitive performance over time, highlighting how repeated exposure to the same tasks created a learning effect:

> **“Remembering the numbers and finding the little diamonds was, again, developing strategies, which sometimes didn’t work. So from that point of view, it was a more of a ‘strategy development’ for me than just a straightforward blank ‘can you remember?’.”**

Participants also found creative solutions to continue with study tasks when experiencing discomfort or when they had concerns around data quality. For example, participants reported using wires, bandages, caps, and gloves over the pulse oximeter or EEG headband to keep them in place, demonstrating how participants attempted to address concerns independently and through experience or trial and error, often without consulting the research team, to continue using the device:

> **“The Velcro pads were useless. I had to secure the headband using wire to connect the pads.”**

#### Theme 4 – Social Support as a Facilitator and a Barrier

“A bit of human contact”

##### Theme Overview

Participants had varying support needs, preferences, and expectations from both the research team and relatives who provided informal study support. Support generally fell into categories: technical support (e.g. to set up mobile applications, enter data, join video calls) and cognitive and emotional support (e.g., reminders on how to complete study tasks, motivation). Having even minimal face-to-face contact with the research team was valued highly and helped participants to continue engagement in the study.

##### Technical Support and Troubleshooting

Someone at home, typically a partner or spouse, provided technical study support for many participants, including some controls who were less familiar or confident with technology. External study support helped participants to navigate complex requirements involved in a study with multiple user interfaces and RMTs. For some participants, this support was limited to the initial set-up such as downloading mobile applications or joining video calls, whilst other participants relied more heavily on support and their partner helped daily, such as inputting digital sleep diary data:

> **“My wife is very technically clued up, and she tends to stand in for me”.**

Participants also contacted the research team for technical support and to report issues such as login failures or software malfunctions. Communication also offered reassurance confirming data transfer and quality and provided a sense of security and connection back to the research team. Regular or available contact helped participants feel supported and engaged during the remote study. Participants reflected specifically on how email communication had an additional benefit of providing a record of the conversation for participants to look back on if they encountered the problem again or had forgotten the advice:

> **“I like using email because I’ve got a record of it.”**

Whilst most participants felt they received sufficient support and training to engage in the remote study tasks, some participants had hoped for more support from the research team, which highlights differences in expectations of support and individual need in remote studies:

> **“A Zoom call, just to quickly refresh, would have been helpful.”**

##### Cognitive and Emotional Support

The involvement of family members not only provided technical assistance, but partners and relatives also helped with cognitive support through reminding participants to complete tasks and helped to answer the subjective questions on sleep:

> **“The main help I had from my wife saying, ‘You know what time, what time do we go to bed?’ And ‘what time do you roughly think we went to sleep?’”**

For some participants, such support was essential for facilitating involvement in the study:

> **“Well, if you interviewed me and my wife, then the answer would be correct, which would be great actually. She provided you with the answers, and she has put the answers in the app.”**

However, several participants, including those with MCI and dementia, did not have any support at home or chose to engage in the study completely independently and reported managing well:

> **“I think I got on OK for myself.”**

Requiring a study partner was identified as a barrier to research participation in other studies. One participant with MCI shared how they had been ineligible for previous research because they do not have someone available to act as a study partner:

> **“I needed to have somebody with me… But yeah, it didn’t, it didn’t come to fruition, because I couldn’t get anybody to join me.”**

##### The Value of Human Contact and Face-to-Face Interactions

Whilst most contact occurred remotely via email or video calls, in-person interactions at the clinic or participants’ homes were valued and helped participants to feel more included in the study:

> **“It’s nice to have a bit of face-to-face and a bit of human contact.”**

Participants also reflected that interacting with researchers frequently and developing rapport helped with confidence and mitigated against study fatigue or disengagement:

> **“Confidence is right. […] Having a short contact with yourself was helpful. I think it helped me.”**

#### Theme 5 – Adherence, Accuracy, and Getting It Right

“I didn’t want to mess it up”

##### Theme Overview

Participants were motivated to complete tasks as they had been instructed to and, for the most part, attempted to adhere to instructions on how and when to complete study tasks to their best of their ability. Participants were sometimes unsure whether devices were working correctly, but reassurance from the research team helped to alleviate concerns and rectify early issues in data collection. Perceptions of suboptimal performance on a task could contribute to negative affect including guilt, frustration, and anxiety.

##### Following the Research Protocol and Wanting to Meet Expectations

Participants took considerable efforts to complete the study tasks as they had been instructed to. When things went wrong or were difficult, some participants experienced negative affect including feeling guilty, embarrassed, or that they had ‘failed’ in some way:

> **“I felt bad because I could not keep the headband on.”**

Knowing that data collection was suboptimal could cause frustration, stress, or disappointment for participants, even when they lacked physical capability to engage in the task:

> **“There didn’t seem to be any saliva at one o’clock in the morning. […] I could happily have spat into test tubes, you know, as much as you wanted. But that wasn’t the case, I know, what you wanted”.**

Being unable to meet study requirements due to conflicting commitments was also stressful for participants, who wanted to follow the study protocol as closely as possible:

> **“It made me feel sort of a little bit kind of guilty, because I’m a conscientious person. […] And then I thought, ‘Oh no, am I ruining the study because I’ve done this late?’”**

Participants also found it more challenging to engage in tasks they subjectively felt they were not performing well on:

> **“You don’t enjoy doing something you’re not very good at.”**

Whilst approaches to and experiences of the cognitive testing differed across participants (**Theme 3**), individuals who felt that they had not performed well or as well as they had expected often experienced frustration or a sense of failure:

> **“It’s very frustrating. There’s nothing about the way you did it, or the methodology, or anything like that. That’s the problem; it’s just that sense of failure because you know you can’t do them.”**

##### Challenges in Accurately Capturing Sleep

Participants felt that recording sleep at home was more comfortable than sleeping in a hospital or sleep laboratory and could provide a more accurate representation of their usual sleep:

> **“All I can say is, if I had had to do this study going into a room in a hospital, or some sort of clinical setting, I cannot imagine getting any sleep at all. So even though the quality of my sleep often wasn’t great, that wasn’t to do with my environment”.**

However, when asked to subjectively describe their sleeping patterns, many participants expressed doubts about the accuracy of their self-reported sleep estimates. Participants often used external cues, such as what television program had been on when they went to bed and what their partner thought, and internal cues such as how rested and awake they felt, to help them estimate nighttime sleep duration, quality, and awakenings:

> **“What I found really, really challenging was being able to hazard a guess about how many hours I’d been awake during the night. Because again, it was just, you know, kind of wild stab in the dark.”**

Participants found it difficult to accurately self-report sleep metrics for their sleep diary, and were aware that their responses might not align with the RMTs. In some cases, writing down sleep patterns appeared to show worse sleep than participants had expected:

> **“You think you sleep better than you actually do.”**

In other cases, trying to complete a sleep diary was challenging to issues with memory recall in addition to being able to estimate duration of time spent awake, either for sleep onset latency or nocturnal awakenings:

> **“Did it take me 5 minutes to go to sleep, or did it take me 30 minutes? It’s all a bit hit and miss.”**

Concerns about devices working as intended and accurately recording sleep data were also prevalent, particularly where participants had subjective sleep disturbance. One participant shared that they were sceptical about whether the devices could reliably track their sleep patterns because of nighttime movements:

> **“I tossed and turned too much. I don’t know whether the headband even managed to gather any data.”**

##### Alleviating Concerns through Reassurance and Clear Instructions

Detailed guidance before completing tasks and opportunities to check data quality for the RMTs helped participants to feel more comfortable and confident with remote data collection. Having printed instructions which were easy to follow helped reassure participants that they were adhering to the research protocols:

> **“I have to re-read it, the instruction, several times just to make sure I was doing it right”.**

Participants often sought reassurance from the research team to check they were meeting study expectations, and appreciated having feedback on whether data collection had been successful, particularly for the EEG headband:

> **“I couldn’t believe it on the first morning after I wore the headset and headband, and got a message from [researcher] saying ‘Wow, it’s come through really clear’.”**

Participants also wanted to know what data was being collected and why, which helped them to feel their efforts had purpose and were being valued. Occasionally, participants were unsure about the rationale behind either the frequency of a task, or the task itself, leading some to question their continued engagement:

> **“I did wonder what I was achieving by doing it”.**

#### Theme 6 – Reflections, Realities, and Uncertainties around Sleep

“Is it just a lazy habit?”

##### Theme Overview

Participants generally perceived good sleep as important for overall and cognitive health but were often unsure how to achieve it. They associated sleep quality with factors such as snoring, dreaming, sleep duration, and nighttime awakenings, and recognised its impact on cognition. Many participants expressed interest in receiving feedback to identify areas for improving their sleep.

##### Factors Influencing Subjective Sleep Quality

Total sleep duration, or getting ‘enough’ sleep, was recognised as a key component of sleep quality. Having approximately eight hours of sleep was recognised as the target for adults, but whether this was always prioritised or perceived as achievable varied considerably:

> **“There’s a lot of talk about ‘you should get sort of eight hours solid sleep a night’ and I don’t know anybody who does”.**
>
> **“I try and get eight hours of sleep at night now. There’s, as a standard for myself”.**

Some participants also felt that adjusting sleep timing by going to bed earlier could be beneficial for sleep, though there was sometimes reluctance towards this, suggesting that a lack of knowledge about good sleep hygiene could contribute towards cognitive dissonance:

> **“Sometimes I think, ‘what’s the point of going to bed early?’ But let’s try and do it.”**

Napping during the daytime was also raised as an important part of sleep among older adults. Some participants were concerned about the health consequences of taking daytime naps, and would actively avoid napping despite daytime sleepiness:

> **“I try to force myself through the day without taking that middle of the afternoon or late afternoon nap.”**

However, others used napping as a strategy to manage energy levels throughout the day or avoid going to bed early:

> **“If I am busy in the day, then when I get home, I do sit in the chair and nap for half an hour.”**

Efforts to adjust sleeping patterns or improve sleep quality largely focused on lifestyle changes and routines which aligned with good sleep hygiene, such as sleep regularity, trying not to sleep during the daytime, avoiding foods which might disturb sleep, and managing stress and emotions:

> **“I’ve got quite a routine anyway. I got to bed about the same time and I go to sleep about the same time and I read for approximately the same amount of time. So, I’m already in a routine”.**

However, participants were keen to seek advice about good sleeping practices, indicating how increased access to knowledge about sleep hygiene could be beneficial among older adults. There were also connotations of getting too much sleep with being lazy or unproductive:

> **“When you wake and you doze, I mean, should you get up and start your day after your five hours? Or is dozing useful? Or is it just a lazy habit?”**

Participants reported how their sleep quality could relate to current circumstances and how stress or worry other co-morbidities could keep them awake at night:

> **“I wake up in the night and can’t get back to sleep. I’m retired, it’s not so bad. But at times, I’ve gone to bed and after an hour I’ve been wide awake because of all of the issues, I’ve been very, very busy at work”.**

Snoring emerged as a major sleep disturbance. Some reported that their snoring was a long-standing issue that significantly affected sleep quality. Snoring affected not only individuals but also their partners, sometimes leading to disrupted sleep, frustration or changes in sleeping arrangements, highlighting the social as well as personal impact of sleep disturbances:

> **“The biggest issue with my sleep has always been my snoring.”**

Vivid dreams were also discussed as a factor affecting sleep quality. These dreams were described as intense and occasionally distressing, with some participants struggling to distinguish between dream content and reality:

> **“I was having quite an intense dream, you know, quite vivid, and it drifted into reality the following day.”**

##### Perceived Consequences of Poor Sleep

Many participants shared how they felt sleep and research investigating how sleep might impact health, cognition, and dementia was important:

> **“The connection between sleep and dementia is important… I was interested to see if my sleep quality affected how I managed the cognitive tests.”**

Some participants noted that they typically felt worse and experienced cognitive impacts after poor sleep, such as experiencing brain fog, difficulty processing information, and lack of motivation:

> **“I’m interested, because I haven’t slept well for so many years, really. And sometimes I feel like I haven’t really slept at all. And it’s very hard to get up and start the day when you’ve had that.”**

However, there was also some doubt about whether sleep had benefits, as they may not have directly noticed a difference in how they felt following sleep:

> **“The only thing for me is that I didn’t perceive any benefit of good sleep”.**

### Comparing Behavioural Drivers and Technology Adoption: Integrating the COM-B Model and UTAUT Framework

We utilised the COM-B model and the UTAUT-2 framework to examine user experience, barriers, and facilitators as both a behaviour more generally (COM-B model) and technology adoption specifically (UTAUT-2).

Our themes and sub-themes mapped to all six components of the COM-B model. *Reflective motivation* related to four of our six themes, highlighting its importance in acceptability (Figure 3). Our themes mapped to all but one component variable in the UTAUT-2 model, *price value*, which was not relevant as study equipment was provided free of charge (Figure 4). We also found potential pathways where lower *age* and greater *experience* might moderate acceptability via influencing familiarity with technology and how challenging it is to use, but no participants referred to *gender* as a relevant moderator.

**Figure 3.**
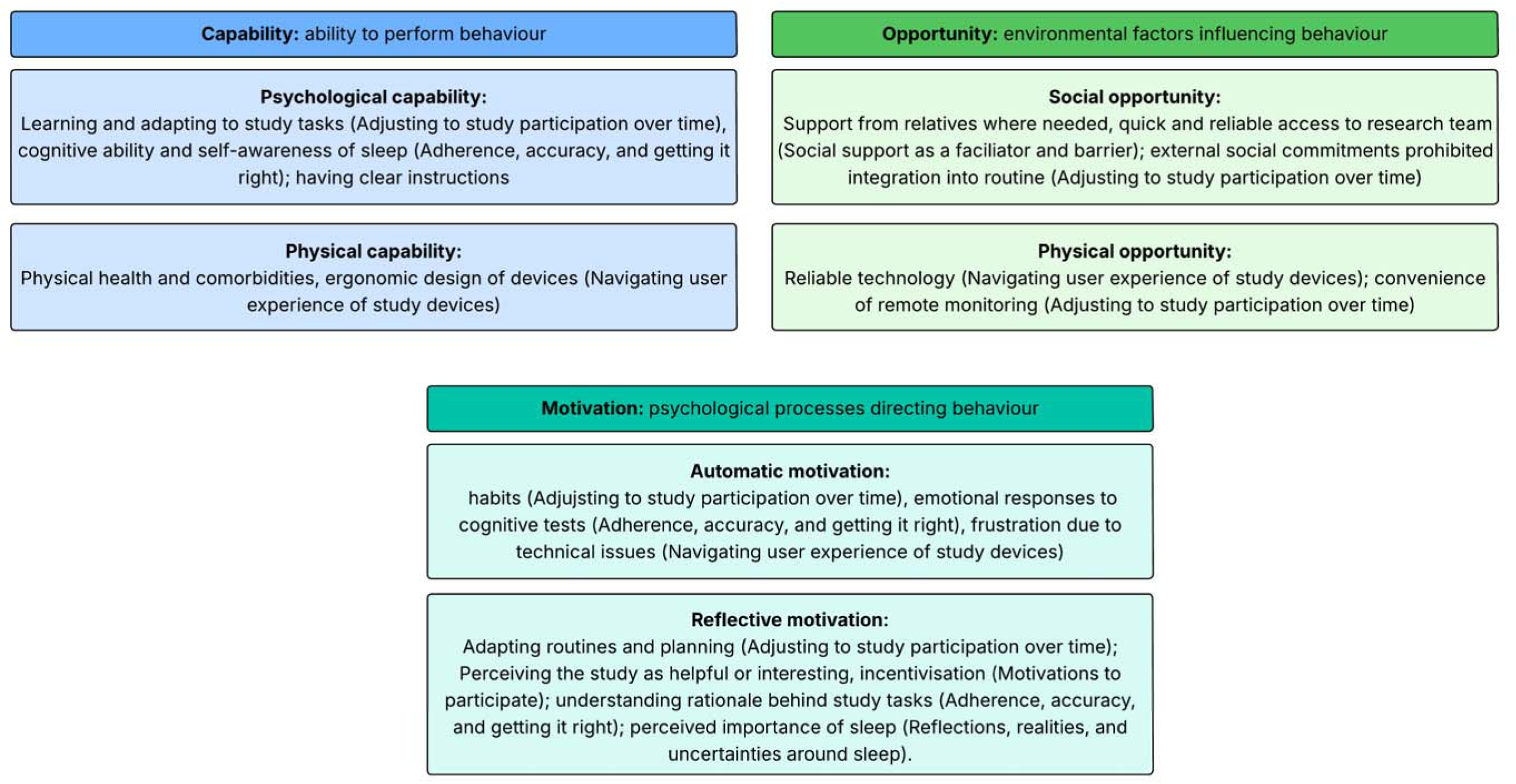
Mapping themes to the Capability, Opportunity, Motivation - Behaviour model of behaviour change.

**Figure 4.**
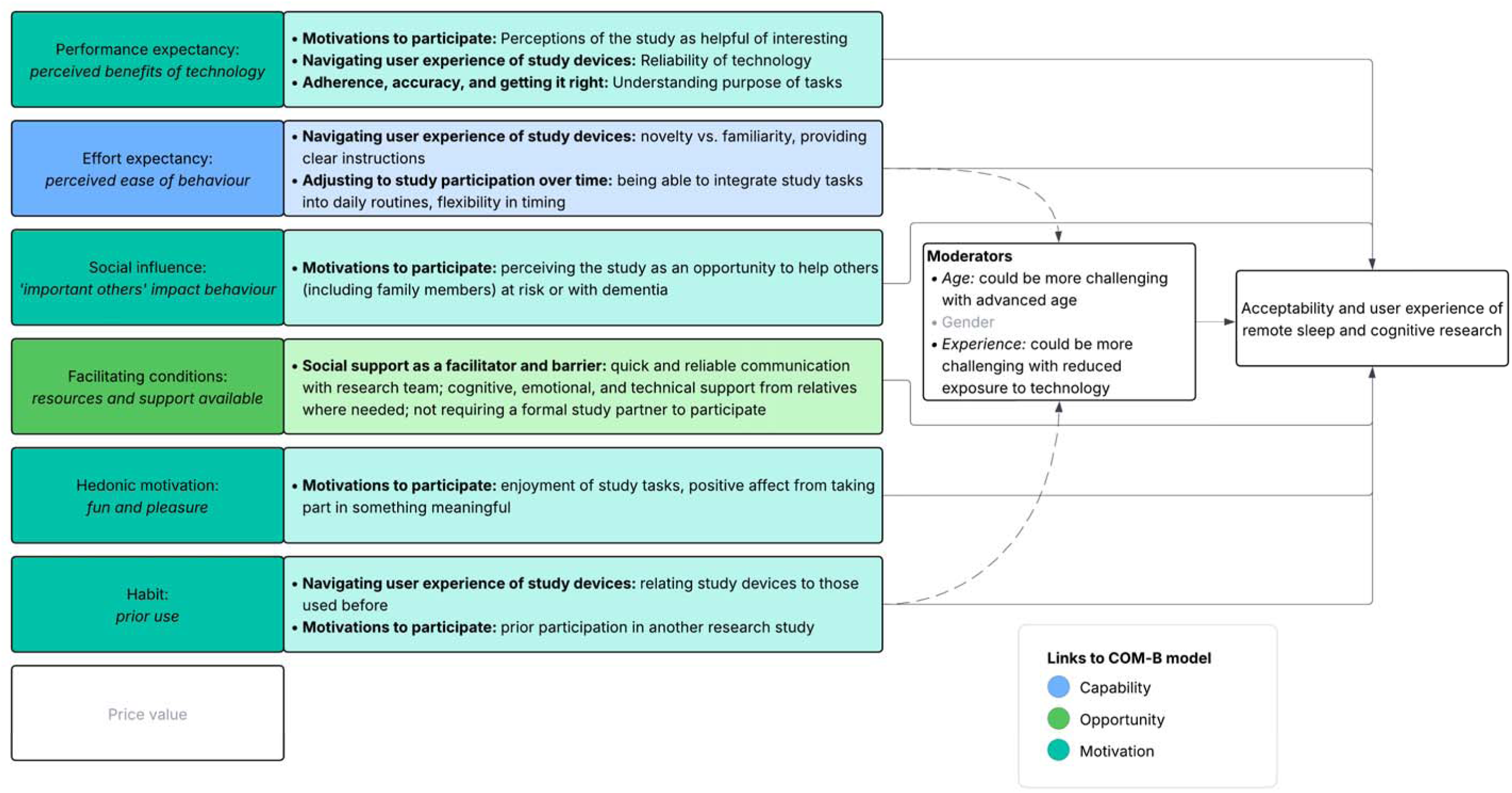
Mapping themes to the extended Unified Theory of Acceptance and Use of Technology (UTAUT-2) model. All concepts

Our findings revealed several intersections between these two models and participants’ experiences. These are explained in detail below for each identified theme.

**Motivations to participate** mapped onto *reflective motivation* (COM-B) as participants were motivated to be helpful and provide meaningful and accurate data. The motivation to be help others also mapped onto *social influence* (UTAUT-2), whilst wanting to provide good data mapped onto *performance expectancy* (UTAUT-2). Study participation provided enjoyment or *hedonic motivation* (UTAUT-2) and participants developed routines or a *habit* (UTAUT-2) to fit in daily tasks, both of which could also come under *automatic motivation* (COM-B).

**Adjusting to study participation over time** through having flexible and convenient study tasks helped participants to have *physical opportunity* (COM-B) to participate and reduced *effort expectancy* (UTAUT-2).

**User experience of the devices** was shaped by *psychological capability* and *physical capability* (COM-B), with digital literacy, physical dexterity, and health comorbidities affecting usability. These aligned with *effort expectancy* and *facilitating conditions* (UTAUT-2), where clear instructions, support from the research team and relatives, and easy-to-use devices were essential for engagement.

**Social support as a facilitator and barrier** mapped onto *social opportunity* (COM-B) because of the importance of support from the research team and relatives, and *social influence* (UTAUT-2), with researcher responsiveness and family assistance increasing confidence and ability to engage in tasks.

**Adherence, accuracy, and getting it right** also mapped onto *reflective motivation* (COM-B) as participants were motivated to provide meaningful and good quality data. However, emotional responses like guilt or anxiety when things went wrong also highlight the psychological toll of tasks which were challenging or where support needs were not met and could be considered to work against *automatic motivation*.

**Reflections, realities, and uncertainties around sleep** also mapped onto *reflective motivation* (COM-B) as participants recognised the importance of sleep, and *performance expectancy* (UTAUT-2) as participants reflected on the accuracy of sleep data collected via sleep diaries and RMTs.

## Discussion

### Principal Findings

Our aim was to explore the views and experiences of older adults with and without cognitive impairment in a research study involving multimodal remote monitoring of sleep and cognition. We identified five themes around barriers and facilitators for remote sleep and dementia research which centred around perceptions of the study as helpful or beneficial (*motivations to participate*), requiring devices to be easy to use, secure, and comfortable (*navigating user experience of devices*), the importance of familiarisation and routine integration (*adjusting to study participation over time*), the value of technical and cognitive support from others and face-to-face human contact (*social support as a facilitator and barrier*), and a drive to ‘do well’ and provide good outcomes (*adherence, accuracy, and getting it right*). Overall, acceptance and engagement were high among older adults with and without cognitive impairment, which aligns with our feasibility findings ^37^. However, we identified several opportunities for improvement in future remote research designs. We also identified a sixth theme, which highlighted interest in sleep hygiene, as well as a knowledge gap, and highlighted the potential utility for sleep education in older adults (*reflections, realities, and uncertainties around sleep*). These themes mapped well onto both the COM-B model (Figure 3) and UTAUT-2 model (Figure 4), suggesting that efforts to improve acceptability and engagement in technology studies and dementia research generally will likely be applicable to remote sleep and dementia research.

Engagement in the study was primarily facilitated by participants’ extrinsic motivations to help others and themselves and intrinsic motivations of interest and enjoyment. Altruism and opportunity for personal benefit are widely cited reasons for research participation and have previously been identified in cognitively healthy older adults engaging in dementia prevention trials ^49^ and patients and caregivers of people living with dementia ^50^. Receiving incentives was also discussed as a potential way to encourage research enrolment or satisfaction, aligning with previous studies in which receiving financial reimbursement for enrolment ^51^, biomarker results ^52^, and feedback on cognitive performance following tasks ^53^ was found to encourage research participation and engagement. In the context of sleep and wearable research, technologies providing high-level personalised sleep summaries for participants or utilising RMTs which automatically provide sleep metrics within an app or onscreen similar to a smartwatch may be worth consideration ^49^.

Convenience and flexibility of study tasks in longitudinal remote research emerged as a key facilitator in allowing research to be integrated into daily routines with minimal burden. Research participation is time-consuming and interfering too much with meaningful activities or previous commitments may be a barrier to research engagement^50^. Remote study tasks help to reduce the time and effort involved in attending clinic visits. However, inflexible scheduling or inconvenient timing of remote data collection may result in missing data or additional participant burden and should be carefully balanced with research aims. For circadian research, it may be desirable or necessary to collect data at inconvenient times, such as around awakening, mealtimes, or bedtime. Our study found that whilst this was acceptable if this was not frequent, greater frequency would likely mean greater burden and reduced acceptability. Researchers should carefully consider which tasks would benefit most from restricted time windows and allow greater flexibility for other variables where feasible. Greater use of passive monitoring devices, which continuously collect data without requiring active involvement from participants, may also help to increase convenience and reduce burden ^54^.

Social support was helpful in encouraging continued participant engagement and fostering positive experiences for some participants, whereas others engaged in the study more independently. These findings highlight the importance of individualised approaches to support. While some participants benefited from or required assistance from relatives for technical and cognitive support, others successfully engaged independently, highlighting that mandatory study partner requirements may unnecessarily exclude capable individuals from research participation. Requiring a study partner has previously been a barrier in similar research studies, particularly for cognitively unimpaired older adults who otherwise meet eligibility criteria ^55^. Despite this, study partners are often considered a requirement for even prodromal dementia research ^55^ ^56^. Study partners in dementia research can offer invaluable support, facilitation, and encouragement, as well as act as informants to help corroborate on symptoms experienced by patients which can be particularly helpful when patients experience difficulties with recall or anosognosia. However, supporting patients with MCI or dementia to engage in research can also have both practical (e.g., logistics of attending visits or calls with researchers) and emotional (e.g., acknowledging cognitive decline) burden on the study partner ^57^, which can prohibit carers, and therefore patients, enrolling in dementia research^50^. For an inclusive and acceptable approach to dementia research, particularly at earlier stages of cognitive impairment, our results highlight the value in flexibility around study partner involvement – benefiting from their support and insight when it is available without excluding patients who would be able to participate independently.

Increasing evidence suggests that individuals with and without cognitive complaints in dementia research studies find remote cognitive testing acceptable ^58^ ^59^, though there remains a preference for human contact. Whilst participants found remote monitoring acceptable, most participants emphasised how regular, real-time remote contact with a researcher was invaluable for troubleshooting, as well as providing opportunities for meaningful social interactions. Occasional face-to-face interactions and timely response to queries were particularly highly valued and helped to overcome technical difficulties as well as sustain engagement and build trust. Automated reminders and written guidance alongside researcher support offered on an *ad hoc* basis was an acceptable level of support for most participants, though some participants would have liked more practical training on technology. Offering regular opportunities to interact with the research team may help to enhance participant engagement and satisfaction, but a flexible approach may be most desirable to avoid participants who do not require the same level of support and may feel “badgered” by too many reminders/follow-ups, possibly increasing attrition ^60^.

Participants evaluated study devices and tasks based on perceptions of comfort achievability, security, and intrusiveness. Technical reliability and simple, integrated platforms were also crucial for maintaining participant confidence and could help to improve data quality. Balancing user comfort and functionality are key features common to ergonomic design ^61^, though RMTs and smart technology focusing on health are often designed to appeal to adults with already healthy lifestyles aiming to optimise and track their habits ^61^, rather than older adults at risk or already with established dementia. Having simple instructions, lower reliance on fine motor skills (such as the use of voice-assisted apps), being minimally intrusive, and balancing familiarity with some variety to maintain interest may be particularly helpful for older adults with cognitive impairment or medical comorbidities. Whilst more interactive study tasks can mean more issues or queries which has led some researchers to encourage moving towards passive RMTs ^55^, our findings suggest that removing all active participant tasks may remove some of the participant satisfaction and engagement. Interactions with RMTs may create opportunities for cognitive stimulation and help foster a sense of meaningful and deliberate contribution to the study ^62^. Balancing passive and active monitoring to ensure sufficient variation to maintain interest and appeal to participants with different preferences may be desirable.

Participants’ user experiences also often changed across the course of the study. Shifts towards positive user experience were observed as participants became more familiar and comfortable with the novel RMTs and successfully integrated tasks into their day by developing routines. However, repetition could be both a barrier and facilitator to continued engagement. For example, whilst some participants gamified the repeated cognitive challenges and saw them as opportunities to improve their scores, for others this repetition was emotionally distressing or fatiguing, particularly if they felt they were not doing well at the task. Most participants continued the tasks in our study ^37^, albeit with reluctance in some cases. In longitudinal ageing studies using repeated cognitive testing, attrition has been linked to lower cognitive and functional ability ^54^. Participants who reported feeling cognitively disengaged may have eventually stopped completing the tasks altogether. Incorporating variation in the study design, such as randomly giving a subset of cognitive tasks each session, as well as reducing the number of tasks which pushed participants to failure, may help to reduce study fatigue and sustain engagement.

Researchers being able to respond quickly to queries or technical problems appeared to be a key facilitator to remote research. Favouring devices which utilise real-time monitoring and feedback may be a way to simultaneously improve participant engagement by establishing a strong connection to the research team, allowing participants to review their own data, and minimising data loss due to technical challenges ^55^. However, whilst many wearable devices and apps offer the benefit of real-time compliance monitoring and feedback ^55^, they typically do not provide context on *why* data is missing. We observed a strong motivation among participants to provide good quality data and providing an opportunity to explain the reason behind missingness (e.g., technical fault needing assistance, conflict with existing commitments, frustration, feeling unwell) may help to identify barriers which can be immediately resolved to reduce further missingness or considered for future studies supporting iterative improvements in remote research design.

### Limitations

The study enrolled participants from one geographical area who were predominantly White British and male, which may limit generalisability to broader, more diverse populations. It is widely understood that more effort is needed to foster diversity in research and ensure equitable access to research ^63^. Remote research utilising various virtual data collection strategies (videoconferencing, online questionnaires, virtual focus groups) to decentralise dementia research may show promise in diversifying trial populations ^64,65^. However, cultural and sex differences may influence the acceptability of research specifically utilising wearable or digital health technologies. For example, individuals who are White and speak English often have more access and are more likely to use digital health technologies ^66,67^, and have fewer concerns about whether research is trustworthy, invasive, or beneficial to their community ^68^. Additionally, men tend to report higher technological confidence ^69^. Lower levels of acceptability or additional barriers may therefore have been identified in a more diverse sample, and research should continue to identify facilitators and barriers across different cultures and demographics.

Self-selection bias may also have influenced the acceptability. Firstly, participants were generally familiar with technology, albeit not most of the devices used in this study. Baseline digital literacy can be a strong predictor of engagement with remote monitoring technologies, with less digitally experienced older adults showing higher dropout rates and reporting greater anxiety when using multiple digital platforms ^70^. It is therefore possible that our findings may not generalise to individuals with lower digital proficiency. Additional support strategies or simplified interfaces may be necessary to ensure equitable access to remote sleep monitoring for all older adults, regardless of their prior technology experience ^71^. Secondly, as only one participant withdrew from the study, and they declined an interview, the analysis focused on participants who engaged throughout the study. Interviewing eligible patients who did not participate in the study, or those who withdrew at an earlier stage, could be invaluable in understanding barriers to participation and what might encourage others to participate ^50^. Future studies should aim to interview caregivers and relatives to corroborate and expand on participants’ experience.

A strength of our study was the duration, as it examined acceptability over eight weeks which allowed participants time to adjust and familiarise themselves with tasks, however our results may not generalise to more extended monitoring periods over many months or years. Given how repeated cognitive testing could lead to fatigue and disengagement, future studies should examine how to optimise acceptability of technology-based research over more prolonged periods in older adults and as cognitive impairment progresses ^70^, as RMTs may be particularly suited to longitudinal cohort studies examining disease progression.

Finally, future research could examine whether there are any systematic differences among individuals at different stages of cognitive impairment in terms of preferences and acceptability, as different levels of engagement can be influenced by cognitive status ^72^. Whilst the quantitative feasibility paper for the RESTED study identified high levels of adherence and good data quality across the AD, LBD, and cognitively healthy controls ^37^, acceptability may differ between cognitively unimpaired vs cognitively impaired older adults.

## Conclusions

Remote monitoring in dementia research offers opportunities to track naturalistic sleep and cognition across disease progression and monitor treatment efficacy in decentralised clinical trials. Incorporating patient-centred methods of remote data collection using predominantly passive RMTs or reliable and easy-to-use apps, combined with periodic face-to-face support, could significantly reduce patient burden, enhance retention, and improve data quality ^55^ in future sleep and dementia research. Our study demonstrates that older adults with MCI and dementia and cognitively unimpaired older adults are willing and able to engage meaningfully with sleep RMTs when provided with appropriate support and flexibility. Key to successful implementation is recognising that acceptability depends on multiple factors, including minimising burden through convenient and flexible task scheduling, ensuring technology is user-friendly and reliable, managing and reducing the burden of challenging or repetitive tasks, and providing tailored and timely support. Understanding the specific barriers and facilitators identified in our study will enable researchers to design more inclusive and effective remote monitoring protocols that accommodate the needs of older adults with cognitive impairment, ultimately advancing our understanding of the relationship between sleep and dementia and supporting future interventional trial design in this area.

## Acknowledgements

The authors sincerely thank the RESTED study participants and their supportive relatives and partners. We also would like to thank the ReMemBr group Lived Experience Experts for their valuable input in refining the study design and enhancing accessibility. We would also like to extend a special thanks to our funders and the teams who supported the project: North Bristol NHS Trust Respiratory Physiology department and the teams at Dreem Research, Cognitron, Join Dementia Research, and Dignio UK.

## Abbreviations

AD: Alzheimer’s disease
LBD: Lewy body disease
MCI: mild cognitive impairment
REM: rapid eye movement
RESTED: Remote Evaluation of Sleep to Enhance Understanding of Early Dementia
RMTs: remote monitoring technologies.

## Data Availability

The data that support the findings of this study are available upon reasonable request from the corresponding author. The data are not publicly available due to privacy or ethical restrictions.

## Authors’ Contributions

BB: Conceptualisation, Methodology, Software, Project administration, Resources, Investigation, Data curation, Formal analysis, Visualisation, Writing – Original draft preparation, Writing – Review & editing.

VGG: Conceptualisation, Methodology, Software, Project administration, Resources, Investigation, Data curation, Formal analysis, Visualisation, Writing – Original draft preparation, Writing – Review & editing.

JB: Conceptualisation, Methodology, Investigation, Resources, Funding acquisition, Writing – Review & editing.

HM: Conceptualisation, Methodology, Investigation, Resources, Funding acquisition, Writing – Review & editing.

EC: Methodology, Funding acquisition, Resources, Supervision, Writing – Review & editing.

AR: Conceptualisation, Methodology, Writing – Review & editing, Supervision.

All authors granted final approval for the version to be published.

## References

1. World Health Organisation. Dementia. Updated 31/03/2025. Accessed 09/04/2025.

2. Gustavsson A, Norton N, Fast T, et al. Global estimates on the number of persons across the Alzheimer’s disease continuum. Alzheimer’s & Dementia. 2023;19(2):658–670. 10.1002/alz.12694

3. Ramanan VK, Day GS. Anti-amyloid therapies for Alzheimer disease: finally, good news for patients. Molecular Neurodegeneration. 2023/06/28 2023;18(1):42. doi:10.1186/s13024-023-00637-0

4. Aljuhani M, Ashraf A, Edison P. Evaluating clinical meaningfulness of anti-β-amyloid therapies amidst amyloid-related imaging abnormalities concern in Alzheimer’s disease. Brain Communications. 2024;6(6)doi:10.1093/braincomms/fcae435

5. Underwood BR. Predicting how many people might receive treatment with new therapies for Alzheimer’s disease. *Journal of Neurology*, Neurosurgery & Psychiatry. 2024;95(9):793–793. doi:10.1136/jnnp-2024-333941

6. Irwin MR, Vitiello MV. Implications of sleep disturbance and inflammation for Alzheimer’s disease dementia. The Lancet Neurology. 2019;18(3):296–306. doi:10.1016/S1474-4422(18)30450-2

7. Simon KC, Cadle C, Shuster AE, Malerba P. Sleep Across the Lifespan: A Neurobehavioral Perspective. Current Sleep Medicine Reports. 2025/02/05 2025;11(1):7. doi:10.1007/s40675-025-00322-2

8. Mc Carthy CE. Sleep Disturbance, Sleep Disorders and Co-Morbidities in the Care of the Older Person. Med Sci (Basel). May 21 2021;9(2)doi:10.3390/medsci9020031

9. Lim MM, Gerstner JR, Holtzman DM. The sleep-wake cycle and Alzheimer’s disease: what do we know? Neurodegener Dis Manag. 2014;4(5):351–62. doi:10.2217/nmt.14.33

10. !!! INVALID CITATION !!! [10];

11. Elder GJ, Lazar AS, Alfonso-Miller P, Taylor JP. Sleep disturbances in Lewy body dementia: A systematic review. Int J Geriatr Psychiatry. Oct 2022;37(10)doi:10.1002/gps.5814

12. Sabia S, Fayosse A, Dumurgier J, et al. Association of sleep duration in middle and old age with incidence of dementia. Nature Communications. 2021/04/20 2021;12(1):2289. doi:10.1038/s41467-021-22354-2

13. !!! INVALID CITATION !!! [12];

14. Eide PK, Vinje V, Pripp AH, Mardal K-A, Ringstad G. Sleep deprivation impairs molecular clearance from the human brain. Brain. 2021;144(3):863–874. doi:10.1093/brain/awaa443

15. Baril AA, Beiser AS, Redline S, et al. Systemic inflammation as a moderator between sleep and incident dementia. Sleep. Feb 12 2021;44(2)doi:10.1093/sleep/zsaa164

16. Maybrier HR, Jackson JJ, Toedebusch CD, Lucey BP, Head D. Influence of sleep and cardiovascular health on cognitive trajectories in older adults. Neurobiology of Aging. 2025/08/01/ 2025;152:34–42. 10.1016/j.neurobiolaging.2025.04.008

17. Bishir M, Bhat A, Essa MM, et al. Sleep Deprivation and Neurological Disorders. Biomed Res Int. 2020;2020:5764017. doi:10.1155/2020/5764017

18. Blackman J, Morrison HD, Lloyd K, et al. The Past, Present and Future of Sleep Measurement in Mild Cognitive Impairment and Early Dementia - Towards a Core Outcome Set: A Scoping Review. Sleep. Apr 4 2022;doi:10.1093/sleep/zsac077

19. Buysse DJ, Hall ML, Strollo PJ, et al. Relationships Between the Pittsburgh Sleep Quality Index (PSQI), Epworth Sleepiness Scale (ESS), and Clinical/Polysomnographic Measures in a Community Sample. Journal of Clinical Sleep Medicine. 2008;04(06):563–571. doi:doi:10.5664/jcsm.27351

20. Buysse DJ, Reynolds CF, Monk TH, Berman SR, Kupfer DJ. The Pittsburgh Sleep Quality Index: a new instrument for psychiatric practice and research. Psychiatry Res. May 1989;28(2):193–213. doi:10.1016/0165-1781(89)90047-4

21. Johns MW. A new method for measuring daytime sleepiness: the Epworth sleepiness scale. Sleep. Dec 1991;14(6):540–5. doi:10.1093/sleep/14.6.540

22. Goparaju B, de Palma G, Bianchi MT. Naturalistic sleep tracking in a longitudinal cohort: how long is long enough? medRxiv. 2024:2024.10.19.24315818. doi:10.1101/2024.10.19.24315818

23. Yoon H, Choi SH. Technologies for sleep monitoring at home: wearables and nearables. Biomed Eng Lett. Aug 2023;13(3):313–327. doi:10.1007/s13534-023-00305-8

24. Reid KJ. Assessment of Circadian Rhythms. Neurol Clin. Aug 2019;37(3):505–526. doi:10.1016/j.ncl.2019.05.001

25. Pullman RE, Roepke SE, Duffy JF. Laboratory validation of an in-home method for assessing circadian phase using dim light melatonin onset (DLMO). Sleep Med. Jun 2012;13(6):703–6. doi:10.1016/j.sleep.2011.11.008

26. Requena-Komuro M-C, Jiang J, Dobson L, et al. Remote versus face-to-face neuropsychological testing for dementia research: a comparative study in people with Alzheimer’s disease, frontotemporal dementia and healthy older individuals. BMJ Open. 2022;12(11):e064576. doi:10.1136/bmjopen-2022-064576

27. Chinner A, Blane J, Lancaster C, Hinds C, Koychev I. Digital technologies for the assessment of cognition: a clinical review. Evid Based Ment Health. May 2018;21(2):67–71. doi:10.1136/eb-2018-102890

28. James CA, Basu T, Nallamothu BK, Kullgren JT. Use of Digital Health Technologies by Older US Adults. JAMA Network Open. 2025;8(1):e2454727–e2454727. doi:10.1001/jamanetworkopen.2024.54727

29. Office for National Statistics. Internet users, UK: 2020. 2021. Accessed 05/06/2025. https://www.ons.gov.uk/businessindustryandtrade/itandinternetindustry/bulletins/internetusers/2020

30. Czaja SJ. Usability of technology for older adults: where are we and where do we need to be. J Usability Studies. 2019;14(2):61–64.

31. Lindqvist E, PerssonVasiliou A, Hwang AS, et al. The contrasting role of technology as both supportive and hindering in the everyday lives of people with mild cognitive deficits: a focus group study. BMC Geriatrics. 2018/08/17 2018;18(1):185. doi:10.1186/s12877-018-0879-z

32. Contreras-Somoza LM, Irazoki E, Toribio-Guzmán JM, et al. Usability and User Experience of Cognitive Intervention Technologies for Elderly People With MCI or Dementia: A Systematic Review. Systematic Review. Frontiers in Psychology. 2021-April-22 2021;Volume 12 – 2021 doi:10.3389/fpsyg.2021.636116

33. Laar A, Silva de Lima AL, Maas BR, Bloem BR, de Vries NM. Successful implementation of technology in the management of Parkinson’s disease: Barriers and facilitators. Clinical Parkinsonism & Related Disorders. 2023/01/01/ 2023;8:100188. 10.1016/j.prdoa.2023.100188

34. Bhargava Y, Baths V. Technology for dementia care: benefits, opportunities and concerns. Journal of Global Health Reports. 2022;6:e2022056.

35. Gabb VG, Blackman J, Morrison HD, et al. Remote Evaluation of Sleep and Circadian Rhythms in Older Adults With Mild Cognitive Impairment and Dementia: Protocol for a Feasibility and Acceptability Mixed Methods Study. Protocol. JMIR Res Protoc. 2024;13:e52652. doi:10.2196/52652

36. Blackman J, Morrison HD, Gabb V, et al. Remote evaluation of sleep to enhance understanding of early dementia due to Alzheimer’s Disease (RESTED-AD): an observational cohort study protocol. BMC Geriatr. Sep 23 2023;23(1):590. doi:10.1186/s12877-023-04288-0

37. Gabb VG, Blackman J, Morrison H, et al. Longitudinal Remote Sleep and Cognitive Research in Older Adults With Mild Cognitive Impairment and Dementia: Prospective Feasibility Cohort Study. JMIR Aging. May 28 2025;8:e72824. doi:10.2196/72824

38. O’Brien BC, Harris IB, Beckman TJ, Reed DA, Cook DA. Standards for Reporting Qualitative Research: A Synthesis of Recommendations. Academic Medicine. 2014;89(9):1245–1251. doi:10.1097/acm.0000000000000388

39. Råheim M, Magnussen LH, Sekse RJ, Lunde Å, Jacobsen T, Blystad A. Researcher-researched relationship in qualitative research: Shifts in positions and researcher vulnerability. Int J Qual Stud Health Well-being. 2016;11:30996. doi:10.3402/qhw.v11.30996

40. Novek S, Wilkinson H. Safe and Inclusive Research Practices for Qualitative Research Involving People with Dementia: A Review of Key Issues and Strategies. Dementia. 2019;18(3):1042–1059. doi:10.1177/1471301217701274

41. Nasreddine ZS, Phillips NA, Bédirian V, et al. The Montreal Cognitive Assessment, MoCA: a brief screening tool for mild cognitive impairment. J Am Geriatr Soc. Apr 2005;53(4):695–9. doi:10.1111/j.1532-5415.2005.53221.x

42. !!! INVALID CITATION !!! ;

43. Ravindran KKG, della Monica C, Atzori G, et al. Evaluation of Dreem headband for sleep staging and EEG spectral analysis in people living with Alzheimer’s and older adults. medRxiv. 2024:2024.12.18.24319240. doi:10.1101/2024.12.18.24319240

44. González DA, Wang D, Pollet E, et al. Performance of the Dreem 2 EEG headband, relative to polysomnography, for assessing sleep in Parkinson’s disease. Sleep Health. 2024/02/01/ 2024;10(1):24–30. 10.1016/j.sleh.2023.11.012

45. Braun V, and Clarke V. Using thematic analysis in psychology. Qualitative Research in Psychology. 2006/01/01 2006;3(2):77-101. doi:10.1191/1478088706qp063oa

46. Proudfoot K. Inductive/Deductive Hybrid Thematic Analysis in Mixed Methods Research. Journal of Mixed Methods Research. 2023;17(3):308–326. doi:10.1177/15586898221126816

47. Venkatesh V, Thong JYL, Xu X. Consumer Acceptance and Use of Information Technology: Extending the Unified Theory of Acceptance and Use of Technology. MIS Quarterly. 2012;36(1):157–178. doi:10.2307/41410412

48. Michie S, van Stralen MM, West R. The behaviour change wheel: A new method for characterising and designing behaviour change interventions. Implementation Science. 2011/04/23 2011;6(1):42. doi:10.1186/1748-5908-6-42

49. Sano M, Egelko S, Zhu CW, et al. Participant satisfaction with dementia prevention research: Results from Home-Based Assessment trial. Alzheimers Dement. Nov 2018;14(11):1397–1405. doi:10.1016/j.jalz.2018.05.016

50. Bouranis N, Gelmon S, Lindauer A. Ability and Willingness to Participate in Dementia Clinical Research: A Qualitative Study. Patient. May 2023;16(3):277–285. doi:10.1007/s40271-023-00621-2

51. McPhillips MV, Petrovsky DV, Brewster GS, et al. Recruiting Persons with Dementia and Caregivers in a Clinical Trial: Dyads Perceptions. Western Journal of Nursing Research. 2022;44(6):557–566. doi:10.1177/01939459211008563

52. Ritchie M, Witbracht M, Russ E, et al. The utility of recruitment incentives in early Alzheimer’s disease trials. Alzheimers Dement. Sep 2024;20(9):6654–6658. doi:10.1002/alz.14143

53. Thompson LI, De Vito AN, Kunicki ZJ, et al. Psychometric and adherence considerations for high-frequency, smartphone-based cognitive screening protocols in older adults. Journal of the International Neuropsychological Society. 2024;30(8):785–793. doi:10.1017/S1355617724000328

54. Popp Z, Low S, Igwe A, et al. Shifting From Active to Passive Monitoring of Alzheimer Disease: The State of the Research. J Am Heart Assoc. Jan 16 2024;13(2):e031247. doi:10.1161/jaha.123.031247

55. Muurling M, de Boer C, Hinds C, et al. Feasibility and usability of remote monitoring in Alzheimer’s disease. DIGITAL HEALTH. 2024;10:20552076241238133. doi:10.1177/20552076241238133

56. Grill JD, Karlawish J. Study partners should be required in preclinical Alzheimer’s disease trials. Alzheimers Res Ther. Dec 6 2017;9(1):93. doi:10.1186/s13195-017-0327-x

57. Black BS, Taylor H, Rabins PV, Karlawish J. Researchers’ perspectives on the role of study partners in dementia research. Int Psychogeriatr. Oct 2014;26(10):1649–1657. doi:10.1017/s1041610214001203

58. Taylor JC, Heuer HW, Clark AL, et al. Feasibility and acceptability of remote smartphone cognitive testing in frontotemporal dementia research. *Alzheimer’s & Dementia: Diagnosis*, Assessment & Disease Monitoring. 2023;15(2):e12423. 10.1002/dad2.12423

59. Gregory S, Harrison J, Herrmann J, et al. Remote data collection speech analysis in people at risk for Alzheimer’s disease dementia: usability and acceptability results. Original Research. Frontiers in Dementia. 2023-October-13 2023;Volume 2 – 2023doi:10.3389/frdem.2023.1271156

60. Teague S, Youssef GJ, Macdonald JA, et al. Retention strategies in longitudinal cohort studies: a systematic review and meta-analysis. BMC Medical Research Methodology. 2018/11/26 2018;18(1):151. doi:10.1186/s12874-018-0586-7

61. Piwek L, Ellis DA, Andrews S, Joinson A. The Rise of Consumer Health Wearables: Promises and Barriers. PLoS Med. Feb 2016;13(2):e1001953. doi:10.1371/journal.pmed.1001953

62. Talbot CV, Briggs P. The use of digital technologies by people with mild-to-moderate dementia during the COVID-19 pandemic: A positive technology perspective. Dementia. 2022;21(4):1363–1380. doi:10.1177/14713012221079477

63. Booi L, Surr C, Greene L, et al. “I am the only one I know of who participates in research”: Promoting Equitable Research Methods for Underserved Communities in Dementia Risk Reduction Research. Alzheimer’s & Dementia. 2023;19(S22):e077944. 10.1002/alz.077944

64. Kim JE, Knox M, Grill JD, et al. Virtual Data Collection Strategies in Research on Alzheimer’s Disease and Related Dementias (ADRD). Innovation in Aging. 2025;9(5)doi:10.1093/geroni/igaf026

65. Yaari R, Holdridge K, Ferguson M, Wessels A, Sims J. Decentralized approaches in TRAILBLAZER-ALZ 3 (P7-6.006). Neurology. 2023;100(17_supplement_2):3010. doi:doi:10.1212/WNL.0000000000202956

66. Woolley KE, Bright D, Ayres T, Morgan F, Little K, Davies AR. Mapping Inequities in Digital Health Technology Within the World Health Organization’s European Region Using PROGRESS PLUS: Scoping Review. J Med Internet Res. Apr 28 2023;25:e44181. doi:10.2196/44181

67. Mitchell UA, Chebli PG, Ruggiero L, Muramatsu N. The Digital Divide in Health-Related Technology Use: The Significance of Race/Ethnicity. The Gerontologist. 2018;59(1):6–14. doi:10.1093/geront/gny138

68. Milani SA, Swain M, Otufowora A, Cottler LB, Striley CW. Willingness to Participate in Health Research Among Community-Dwelling Middle-Aged and Older Adults: Does Race/Ethnicity Matter? J Racial Ethn Health Disparities. Jun 2021;8(3):773–782. doi:10.1007/s40615-020-00839-y

69. Christensen MA. Tracing the Gender Confidence Gap in Computing: A Cross-National Meta-Analysis of Gender Differences in Self-Assessed Technological Ability. Social Science Research. 2023/03/01/ 2023;111:102853. 10.1016/j.ssresearch.2023.102853

70. Kebede AS, Ozolins L-L, Holst H, Galvin K. Digital Engagement of Older Adults: Scoping Review. Review. J Med Internet Res. 2022;24(12):e40192. doi:10.2196/40192

71. Tsai HS, Shillair R, Cotten SR. Social Support and “Playing Around”: An Examination of How Older Adults Acquire Digital Literacy With Tablet Computers. J Appl Gerontol. Jan 2017;36(1):29–55. doi:10.1177/0733464815609440

72. Holthe T, Halvorsrud L, Karterud D, Hoel KA, Lund A. Usability and acceptability of technology for community-dwelling older adults with mild cognitive impairment and dementia: a systematic literature review. Clin Interv Aging. 2018;13:863–886. doi:10.2147/cia.S154717

