## Supplementary Files for "“Like taking part in Star Wars”: A thematic analysis of acceptability and experiences of older adults participating in remote longitudinal sleep and dementia research"

Supplementary Materials

### I. Standards for Reporting Qualitative Research (SRQR) Checklist

Adapted from: O’Brien B.C., Harris, I.B., Beckman, T.J., Reed, D.A., & Cook, D.A. (2014). Standards for reporting qualitative research: a synthesis of recommendations. Academic Medicine, 89(9), 1245-1251.

| No. Topic | Item | Location of evidence |
| --- | --- | --- |
| **Title and abstract** | | |
| S1 Title | Concise description of the nature and topic of the study identifying the study as qualitative or indicating the approach (e.g., ethnography, grounded theory) or data collection methods (e.g., interview, focus group) is recommended | Title of paper |
| S2 Abstract | Summary of key elements of the study using the abstract format of the intended publication; typically includes objective, methods, results, and conclusions | Structured abstract |
| **Introduction** | | |
| S3 Problem formulation | Description and significance of the problem/phenomenon studied; review of relevant theory and empirical work; problem statement | Introduction: ‘Sleep, aging, and dementia’ and ‘Measuring sleep in individuals with and at risk of dementia’ |
| S4 Purpose or research question | Purpose of the study and specific objectives or questions | Introduction: ‘Aims of the present study’ |
| **Methods** | | |
| S5 Qualitative approach and research paradigm | Qualitative approach (e.g., ethnography, grounded theory, case study, phenomenology, narrative research) and guiding theory if appropriate; identifying the research paradigm (e.g., positivist, constructivist/interpretivist) is also recommended | Methods: ‘Data analysis’ – “A combination of deductive and inductive thematic analysis was conducted” |
| S6 Researcher characteristics and reflexivity | Researchers’ characteristics that may influence the research, including personal attributes, qualifications/experience, relationship with participants, assumptions, or presuppositions; potential or actual interaction between researchers’ characteristics and the research questions, approach, methods, results, or transferability | Methods: ‘Reflexivity’ |
| S7 Context | Setting/site and salient contextual factors; rationale | Methods: ‘Reflexivity’, ‘Study design’ |
| S8 Sampling strategy | How and why research participants, documents, or events were selected; criteria for deciding when no further sampling was necessary (e.g., sampling saturation); rationale | Methods: ‘Participants’ |
| S9 Ethical issues pertaining to human subjects | Documentation of approval by an appropriate ethics review board and participant consent, or explanation for lack thereof; other confidentiality and data security issues | Methods: ‘Ethical considerations’ |
| S10 Data collection methods | Types of data collected; details of data collection procedures including (as appropriate) start and stop dates of data collection and analysis, iterative process, triangulation of sources/methods, and modification of procedures in response to evolving study findings; rationale | Methods: ‘Study design’, Figure 1 |
| S11 Data collection instruments and technologies | Description of instruments (e.g., interview guides, questionnaires) and devices (e.g., audio recorders) used for data collection; if/how the instrument(s) changed over the course of the study | Supplementary Materials |
| S12 Units of study | Number and relevant characteristics of participants, documents, or events included in the study; level of participation (could be reported in results) | Methods: ‘Participants’ |
| S13 Data processing | Methods for processing data prior to and during analysis, including transcription, data entry, data management and security, verification of data integrity, data coding, and anonymization/deidentification of excerpts | Methods: ‘Data analysis’ |
| S14 Data analysis | Process by which inferences, themes, etc., were identified and developed, including researchers involved in data analysis; usually references a specific paradigm or approach; rationale | Methods: ‘Data analysis’ |
| S15 Techniques to enhance trustworthiness | Techniques to enhance trustworthiness and credibility of data analysis (e.g., member checking, audit trail, triangulation); rationale | Methods: ‘Data analysis’ |
| **Results/Findings** |  |  |
| S16 Synthesis and interpretation | Main findings (e.g., interpretations, inferences, and themes); might include development of a theory or model, or integration with prior research or theory | Results: ‘Summary of themes’ and ‘Comparing Behavioural Drivers and Technology Adoption: Integrating the COM-B Model and UTAUT Framework’ |
| S17 Links to empirical data | Evidence (e.g., quotes, field notes, text excerpts, photographs) to substantiate analytic findings | Results |
| **Discussion** |  |  |
| S18 Integration with prior work, implications, transferability, and contribution(s) to the field | Short summary of main findings; explanation of how findings and conclusions connect to, support, elaborate on, or challenge conclusions of earlier scholarship; discussion of scope of application/generalizability; identification of unique contribution(s) to scholarship in a discipline or field | Discussion: ‘Principal Findings’ |
| S19 Limitations | Trustworthiness and limitations of findings | Discussion: ‘Limitations’ |
| **Other** |  |  |
| S20 Conflicts of interest | Potential sources of influence or perceived influence on study conduct and conclusions; how these were managed | Conflicts of interest |
| S21 Funding | Sources of funding and other support; role of funders in data collection, interpretation, and reporting | Funding sources |

### II. Interview topic guide

**Remote Evaluation of Sleep To enhance understanding of Early Dementia (RESTED)**

**Semi-Structured Interview Topic Guide**

The semi-structured interview as part of the RESTED study is designed to gain valuable insight from a range of participants who may have completed or withdrawn from the study. This interview schedule has been designed with fields of implementation science and health psychology in mind and draws off elements from the COM-B (Capability, Opportunity, Motivation and Behaviour) Model.

The overall purpose of the interview is to explore two major themes:

1. Participant experience of interacting with and using supplied technological equipment.
2. Participant experience of involvement in a study primarily designed to be delivered remotely.

**Guide for Overall Interview Structure**

**Part One – Welcome**

To include welcome, introduction, explanation of interview plan (including expected time and purpose) and our thanks for participation / time commitment.

**Part Two – Introductory Questions**

- “How did you first hear about the study?”
- “Why did you decide to take part?”
- “Was there anything that you were worried about before starting the study?”
  - About your health
  - About what the study might ask you to do
  - About being able to use the technology/equipment
  - About your friends/family/carers (helping or hindering)

**Part Three – Study Equipment**

To explore experiences of the following:

- Dignio Application via Portable Device
- Study Prompts via Portable Device
- Short Cognitive Tests via Cognitron
- Cognitive Word List Tasks via Cognitron / Telephone
- Recording Sleep Diary Entries
- Autobiographical Memory Task
- Wearing Wrist Actigraph
- Wearing and Operating Dreem EEG Headband
- Supplying Saliva Swabs

For each of the above stems may include:

- “Tell me what it was like…. (e.g.) wearing the watch?”
- “What was the worst or most annoying thing about (e.g.) wearing the watch?”
- “Was there anything that you thought was good / novel / interesting about … (e.g.) the short cognitive tests?”
- “Was there anything that was particularly difficult about… (e.g.) wearing the watch”
- “Do you think that being involved in RESTED changed your sleep, or your attitudes towards sleep, in any way?”

**Part Four – Remote vs Face-to-Face**

- “Could you tell us what you thought about having mostly remote, by telephone etc, rather than face-to-face discussions with the study team?”

Also, to explore individual modalities of remote contact including telephone and online video consultation.

- “Could you tell us how you most like to have contact with the research team?
  - If you couldn’t meet face-to-face, do you prefer telephone or online contact?
  - And why?
- “Was there anything you particularly liked about… e.g., telephone contact?”
- “What was challenging or more difficult about… e.g., telephone contact?”
- “How did you feel about completing the surveys via an app, as opposed to completing paper forms or using a website?”

**Part Five – Overall impression of the study and remote sleep technology and research**

- “We are interested in your overall impression and thoughts about being involved in the study. While we would really like to hear about things that went well, we are particularly interested in anything which could have been done better or that was more difficult for any reason. This is so that we can learn the most from what we have done and also influence future research for the better.”
- “With this in mind, could you tell us whether there was anything that you did not like about the study?”
  - What didn’t work for you?
  - What made it hard to take part?
  - Was there anything that was frustrating or annoying, or just a pain to have to do?
- “What (if anything) was difficult or challenging about the study”?
- “What could we do to make it easier for other people to take part in the study if we did it again?
- **“**What went well in the study?” or “Was there anything that you found particularly enjoyable as part of the study?”
- “Is there anything you expected us to measure (which we didn’t) with regards to your sleep?”

“We would like to now ask you some more general questions about sleep technologies to help guide future research.”

- “For the purposes of RESTED, we don’t feedback to patients about their sleep data, but future sleep technology or research could offer this. Would you want access to sleep data, and do you think sleep data could be useful to you? In what ways?”
- “What do you think about others (e.g., family members, carers, healthcare professionals) having access to your sleep data?”
- “Did you ask for help on the study tasks from someone not in the research team (e.g., family member, carer, or friend)? Do you think taking part in RESTED affected your relationships with others?
- “Is there anything you expected us to ask you about?”
- “Is there anything else which you feel you wanted to say that you haven’t had the opportunity to get across?”

**Part Six – Concluding Remarks**

To ask the participant if they have any questions and to include our thanks for participation in the both the whole study and this interview.

### III. Questionnaire on motivations, expectations, and experience with the technology

Note: Responses were provided in free-text unless otherwise described. This questionnaire was administered via the *MyDignio* mobile app immediately following baseline assessments.

We would like to find out a little more about how you feel about taking part in the study, and your previous experiences with technology.

**1. Why did you decide to take part in the RESTED study?**

**2. How do you feel about taking part in the RESTED study?**

**3. What are your first impressions of the sleep monitoring devices (e.g., actigraphy watch, Dreem headband) you’ll be using in RESTED?**

**4. How confident do you feel about using the Dignio app to answer questions on your sleep?**

Very confident / Somewhat confident / Not very confident / Not at all confident / I’m not sure

**5. How often do you typically use a smartphone, tablet, computer, or smart watch in your day-to-day life?**

Multiple times per day / At least once per day / Most days / Some days / Rarely / Not at all

**6. Have you used any of the technology that is used in RESTED before (e.g., an overnight pulse oximeter, an actigraphy watch)?**

Yes / No / Not sure

**7. Have you used any other ‘wearable’ devices to monitor your sleep (e.g., a smart watch)?**

Yes / No / Not sure

**8. How have your family members/friends reacted regarding you participating in a remote study for sleep research?**

### IV. Questionnaire on study experiences so far and intensive week (“Intensive week experiences week questionnaire”)

Note: Responses were provided in free-text unless otherwise described. This questionnaire was administered via the *MyDignio* mobile app immediately following baseline assessments.

We would like to find out a little more about how you felt about the ‘intensive week’ of the RESTED study (when you wore the Dreem headband and did additional study tasks).

**1. Overall, in a sentence or two, how did you find the ‘intensive week’ of the RESTED study?**

**2. Which ‘intensive week only’ study task(s) did you find the easiest to complete? [select as many as you want]**

Providing saliva samples / Daily cognitive tests via Cognitron / Memory tests with the research team / Wearing the Dreem Headband overnight

**3. Did you find any of the ‘intensive week’ study tasks particularly difficult to complete? [select as many as you want]**

Providing saliva samples / Daily cognitive tests via Cognitron / Memory tests with the research team / Wearing the Dreem Headband overnight

**4. Is there anything that you think would have made the ‘intensive week’ of RESTED easier or better for you?**

**5. How have you felt about completing the remote cognitive tests via Cognitron daily during the intensive week?**

**6. How easy or difficult has it been to remember and have time to complete the daily sleep diary? [select one]**

Very easy / Quite easy / Neither easy nor difficult / Quite difficult / Very difficult

**7. Have you noticed any changes (positive or negative) in your relationship with your partner or others for using the sleep technologies?**

Yes / No

**If yes, please explain.**

**8. Do you think taking part in the RESTED study has changed any of your daily activity or sleep habits?**

Yes / No

**If yes, please explain:**

**9. Do you have any other feedback for us at this point?**
